# The impact of morbidity burden on cancer diagnosis; a retrospective cohort study in England

**DOI:** 10.1101/2025.08.28.25334632

**Authors:** Bianca Wiering, Luke TA Mounce, Sarah J Price, David Shotter, Jose M Valderas, Samuel WD Merriel, Sarah Moore, Leon Farmer, Christian Von Wagner, Rupert A Payne, Cristina Renzi, Georgios Lyratzopoulos, Willie Hamilton, Gary A Abel

## Abstract

**Background:** Expediting cancer diagnosis is a priority in many countries. The rising prevalence of chronic conditions may complicate the cancer diagnostic process. We investigated whether patients with pre-existing morbidity were more likely to experience disadvantage in cancer diagnostic outcomes and processes.

**Methods:** We used linked primary, secondary care, and cancer registration data for patients aged 40+ years diagnosed with incident cancer in England during 2012-2018. The Cambridge Multimorbidity Score quantified morbidity burden. Logistic regressions investigated whether morbidity burden was associated with stage at diagnosis, 30-day all-cause mortality, emergency presentation- or urgent suspected cancer referral route to diagnosis.

**Results:** 288,297 patients were included. Decreasing morbidity burden was associated with an increased likelihood of advanced-stage diagnosis (e.g. high burden vs. no burden aOR: 0.72, 95% CI: 0.7-0.75, p<0.0001). There were u-shaped relationships between morbidity burden, emergency diagnoses and 30-day mortality, with those with high or no morbidity burden most likely to be diagnosed as an emergency and to die within 30 days after diagnosis. Diagnoses via urgent suspected cancer referrals decreased with increasing morbidity burden. Associations varied across cancer sites, though higher morbidity burden was not associated with advanced stage for any cancer.

**Conclusion:** Contrary to expectations, not having pre-existing morbidities was associated with an increased risk of advanced-stage diagnosis and emergency presentations. This may reflect heightened surveillance of patients with morbidity being protective against later advanced-stage cancer diagnoses. These findings highlight the need for robust cancer surveillance processes and good comprehensive care that considers cancer alongside wider aspects of health.

## Introduction

Survival of cancer patients varies across countries [1, 2], which may in part be due to differences in pathways and intervals to diagnosis [3]. Early-stage cancer diagnosis benefits both patients and health care systems. It increases the number of available treatment options and their success, and reduces treatment length, costs, complications, and long-term impacts of the cancer and its treatment [2, 4–6]. Whilst there have been improvements in earlier diagnosis and survival [1, 3, 7], the prevalence of morbidity has also increased [8, 9]. It is estimated that more than a third of adults worldwide have two or more long-term-conditions [10]. The degree of morbidity increases with age and age is the predominant risk-factor for cancer [11]. For cancer patients, current evidence present a conflicting picture of the relationship between pre-existing morbidities, patient outcomes and cancer diagnostic timeliness [12]. A recent review found that generally pre-existing morbidities are associated with longer diagnostic timelines, a lower likelihood of receiving standard care, more complications and reduced survival [13]. The evidence on stage at diagnosis was more mixed, with slightly more studies favouring a negative impact. Many of the included studies, however, were focused on single morbidities and/or single cancer sites.

Three principal mechanisms have been proposed by which morbidity could interact with the cancer diagnostic process [13]. In England, most patients are diagnosed with cancer following presentation in primary care [14], where action is taken based on symptoms, signs, or test results. Symptoms, and care processes relating to pre-existing morbidities – and their shared risk factors, such as age and socioeconomic deprivation – may interfere with clinical decision making [11, 15]. The first mechanism operates when a pre-existing morbidity offers a plausible ‘alternative explanation’ for cancer features [16, 17]. Features may be misattributed to the morbidity or its treatment, instead of patients and/or clinicians considering cancer. The nature of the morbidity plays a significant role in this scenario. Second (‘competing demands’), symptoms are recognised as possible cancer symptoms; however, with limited time, resources and ongoing morbidity management, cancer investigations may have to compete with morbidity management demands for clinical care [18]. While the nature of the morbidity may play a role, the accumulated morbidity burden across conditions may be more important here. Third, regular healthcare encounters for morbidity management may facilitate cancer identification, as it allows for more opportunities to discuss and investigate cancer symptoms – a ‘surveillance’ mechanism. Additionally, tests or scans conducted for monitoring morbidities could also help detect cancer [19–21]. Both the nature of the morbidity and morbidity burden are likely important here.

No comprehensive study exploring whether pre-existing morbidity may affect patient outcomes and routes to diagnosis for all cancers using one pre-defined method has yet been conducted. As morbidity burden may be important for several mechanisms, here, we aimed to investigate whether patients with increasing morbidity burden were more likely to be disadvantaged in terms of cancer diagnostic outcomes and routes to diagnosis. More specifically, our primary research question was:

1. Are patients with higher morbidity burden more likely to be diagnosed with advanced-stage cancer? The secondary research questions were:
2. Are patients with higher morbidity burden more likely to be diagnosed with cancer after an emergency presentation, or by urgent suspected cancer referral?
3. Are patients with higher morbidity burden more likely to die within 30 days of diagnosis?

## Methods

### Data sources

The Clinical Practice Research Datalink (CPRD) Aurum is a validated database consisting of de-identified primary care health records routinely collected from a network of UK GP practices. CPRD Aurum covered 10% of GP practices in England in 2018 [22, 23]. For this study, CPRD Aurum data relating to English patients were linked to the National Cancer Registration and Analysis Service (NCRAS) [24], Hospital Episode Statistics (HES) [25], patient-level Index of Multiple Deprivation (IMD) 2015, and Office for National Statistics (ONS) death data [22, 26].

### Study population

We included patients with a record of incident cancer in NCRAS between 01/01/2012 and 31/12/2018 with data available in CPRD. Patients had to be 40 years or older at the time of diagnosis with at least three years’ registration at their general practice before diagnosis. Patients who were diagnosed via screening, were diagnosed with non-melanoma skin cancer, had a diagnosis recorded after their death date, had a recorded gender not matching a sex-related cancer, or had a cancer diagnosis recorded before the inclusion period were excluded.

### Ethical approval

The National Research Ethics Service (NRES) has granted ethical approval for observational research using anonymised CPRD data. Observational studies using anonymised CPRD data are therefore not required to obtain study-specific ethical approval [27]. Study protocol approval was granted by the CPRD Independent Scientific Advisory Committee (protocol 20_126R).

### Patient and public involvement (PPI)

The study is embedded in a NIHR funded programme grant (SPOCC). Extensive PPI consultation was conducted in the design of SPOCC. SPOCC has a patient co-investigator, and an overarching PPI group, with individual members taking bridge roles, attending project and PPI meetings. For this study, the PPI bridge (LF) attended three project meetings, and one meeting was held to disseminate results to the PPI group. LF and the PPI-co-investigator attended programme meetings; LF and work package leads were in regular contact; and LF is a co-author for this manuscript.

### Cancer site and stage

Cancer site and year of diagnosis were extracted from NCRAS. If there were multiple diagnoses recorded on the same date, the highest staged cancer was retained. For the few remaining multiple diagnoses, a diagnosis was selected at random. Based on ICD-10 codes (C00-C97 and D05.1, but not C44), 25 common sites were identified plus carcinoma in situ of breast, with remaining cancers assigned to an “other” category (Appendix A: Table A.1).

Where available, cancer stage was extracted from NCRAS using the TNM system (Appendix A: Table A.2 for missing stage information). Cancer stage was divided into two groups. Group one (early stage) included stages 0, I and II, and group two (advanced stage) included stages III and IV. Cancer sites which are not stageable in the TNM system were excluded from stage analyses.

### Mortality and route to diagnosis

The derived route to diagnosis, which includes two-week-wait referral (urgent referral for suspected cancer) and emergency presentation, among others, was extracted from NCRAS [28]. We used this information to create two binary variables, one indicating whether patients were diagnosed after an urgent suspected cancer referral, and the other indicating whether patients were diagnosed after an emergency presentation (i.e. emergency referral, transfer, admission, or attendance to or within secondary care). The date of death, as recorded in ONS data, was used to create a binary variable indicating whether patients had died from any cause within 30 days after their cancer diagnosis.

### Morbidity burden

Morbidity burden was the explanatory variable in all main analyses and was defined as the overall impact of a range of morbidities present at least a year before the cancer diagnosis. Morbidity burden was estimated using the Cambridge Multimorbidity Score (CMS) general outcome weighting [29]. The CMS has been validated and shown to outperform the Charlson Comorbidity Index. Furthermore, the CMS covers more conditions, and is weighted by primary care use, unplanned hospital admissions and mortality. The CMS makes use of medical and prescription codes recorded in CPRD for 37 conditions, and has requirements for the number, type, and time of recording of codes per morbidity [29]. Where possible, pre-existing validated code lists were used. If no suitable code list existed, robust methods were used to collate and validate SNOMED-CT codes that GPs might use [30]. The CMS was used to create four morbidity burden groups (none, low, medium, high), with the last three groups based on CMS tertiles.

### Patient characteristics

Age (5-year groups between 40 and 89 years and a 90 years and older group), gender, and smoking history (never/ever) were extracted from CPRD. National quintiles were used to define IMD groups.

### Data analysis

Separate logistic regression models were used to investigate whether patients with higher morbidity burden were more likely to be diagnosed with advanced stage (vs early stage), to be diagnosed after an emergency presentation (vs other route) or urgent suspected cancer referral (vs other route), or to die within 30 days of their cancer diagnosis. All main effects-only models included morbidity burden and cancer site as exposure variables, and patient age, gender, deprivation, year of diagnosis, and smoking history as covariates. Standard errors were adjusted for clustering of observations at general practice-level. Subsequently, 2-way interaction terms between cancer site, patient characteristics and morbidity burden were investigated individually with all interactions retained in the models. Significance was tested with a joint Wald test. Sensitivity analyses included a composite outcome combining advanced stage with emergency route to diagnosis, the addition of stage as a covariate for 30-day mortality and routes to diagnosis models, and two alternative morbidity measures, a morbidity count, and the Johns Hopkins Adjusted Clinical Groups (ACG) system [31] (Appendix A). Analyses were conducted using Stata v.17 [32]. Figures 1 to 4 were produced using R [33].

**Figure. 1.**
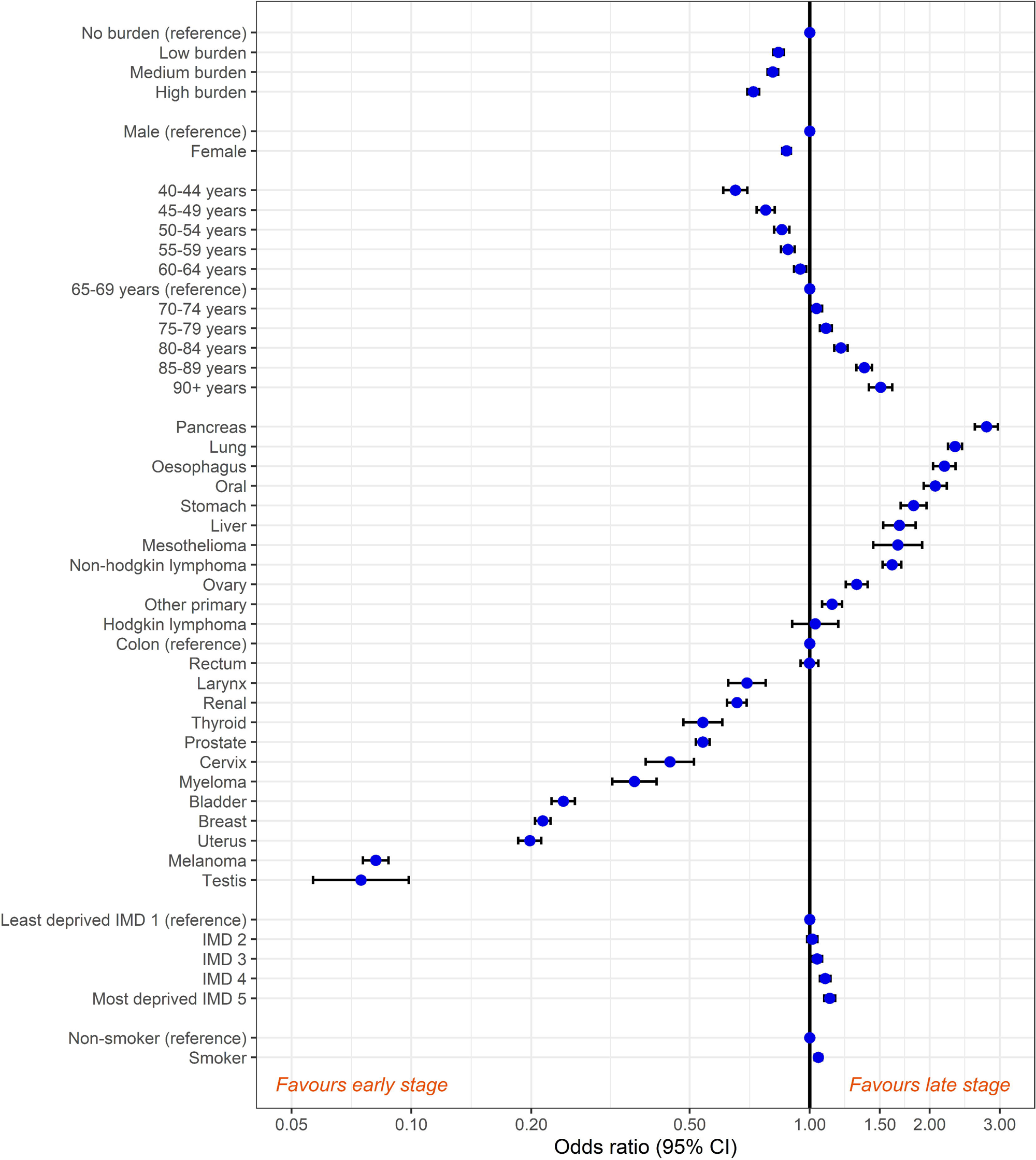
Associations between cancer stage, patient characteristics, cancer site and morbidity burden*. *Confidence intervals were provided for all associations. However, narrow intervals may not be visible as they overlap with the dot indicating the odd ratio point estimate.

### Funding source

This study is funded by the NIHR Programme Grants for Applied Research (PGfAR) SPOtting Cancer among Comorbidities (SPOCC) programme: supporting clinical decision making in patients with symptoms of cancer and pre-existing conditions (NIHR201070). The funder had no role in study design, data collection, analysis, interpretations, or writing of the report.

## Results

### Patient characteristics

We included 288,297 patients meeting the inclusion criteria. Of these, 154,614 (53.6%) patients were male, and the mean age was 70 years (Range: 40 - 107.8; SD: 12.5) (Table 1). The most common cancers were prostate (n= 46,631; 16.2%) and lung (n= 38,189; 13.3%). Most patients had at least one pre-existing morbidity (n=241,677; 83.8%). The mean Cambridge Multimorbidity Score was 1.3 (Range: −0.02 – 11.6; SD: 1.4). 24,335 patients died (8.4%) within 30 days of their diagnosis and 110,450 out of 220,617 (50.1%) of staged patients were diagnosed with advanced-stage cancer. Of all patients, 117,306 (40.7%) were diagnosed after an urgent suspected cancer referral and 58,649 (20.3%) as an emergency (Table 2).

**Table 1.**
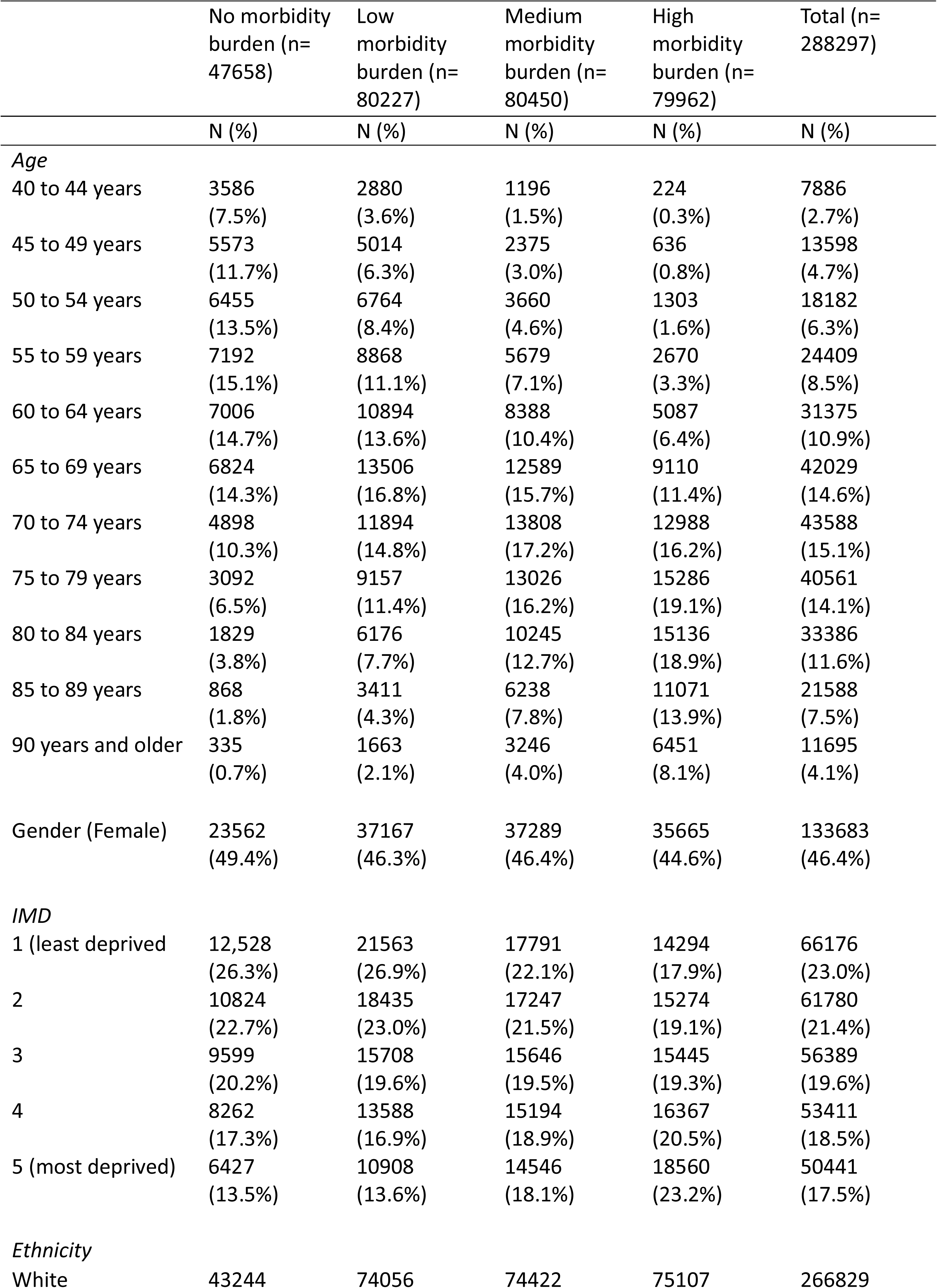
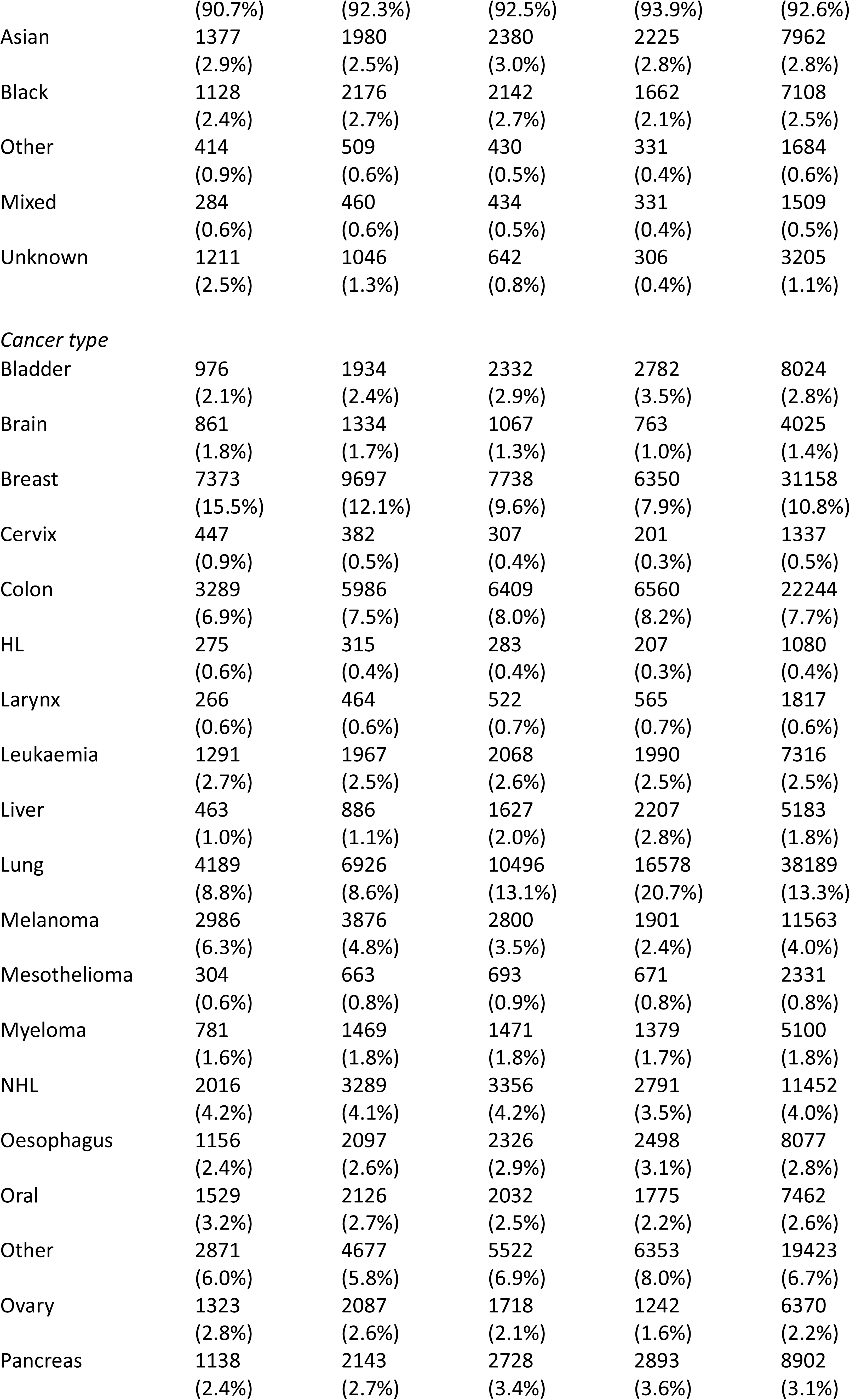
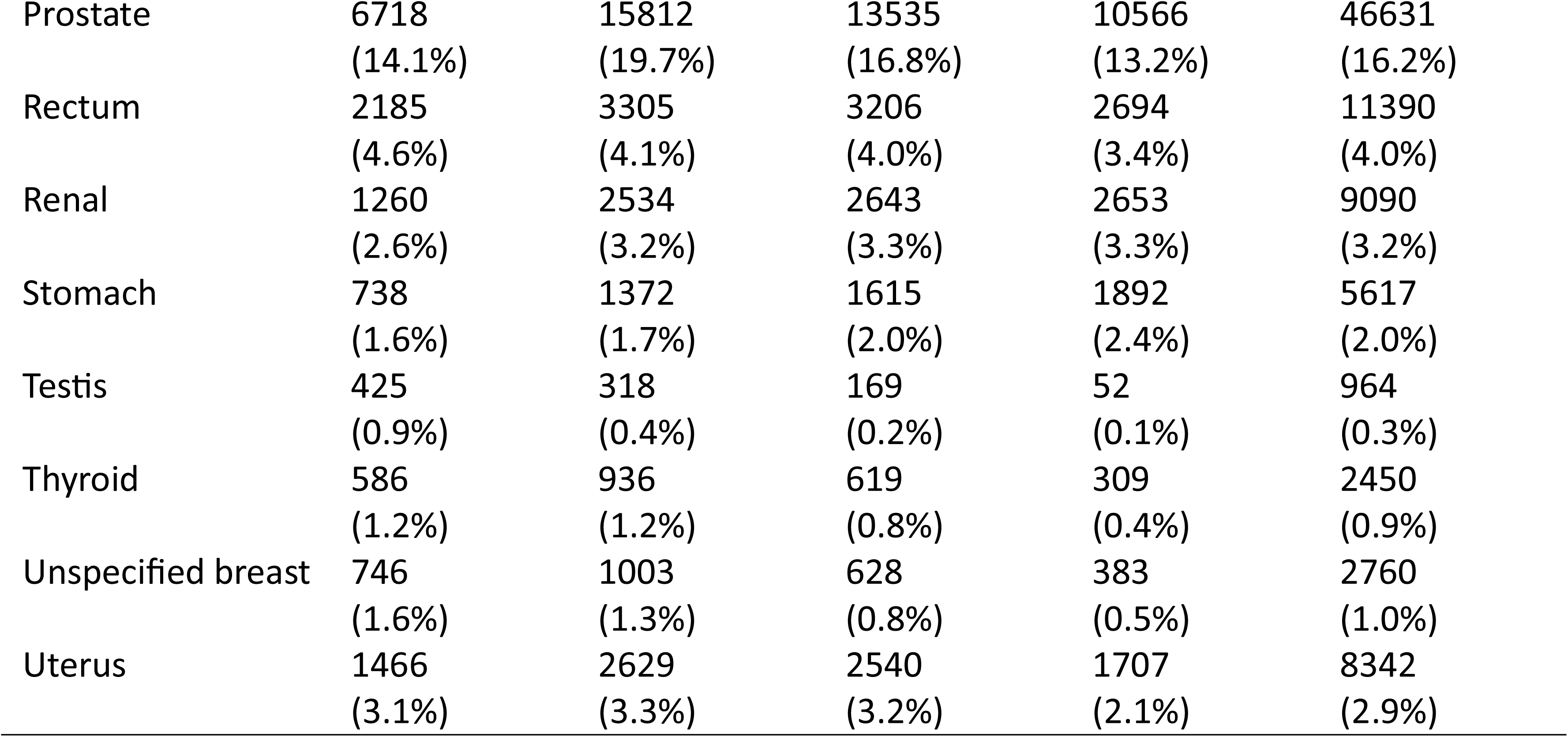
Patient characteristics by morbidity burden grouping.

|  | No morbidity<br>burden (n=<br>47658) | Low<br>morbidity<br>burden (n=<br>80227) | Medium<br>morbidity<br>burden (n=<br>80450) | High<br>morbidity<br>burden (n=<br>79962) | Total (n=<br>288297) |
| --- | --- | --- | --- | --- | --- |
|  | N (%) | N (%) | N (%) | N (%) | N (%) |
| *Age* |  |  |  |  |  |
| 40 to 44 years | 3586<br>(7.5%) | 2880<br>(3.6%) | 1196<br>(1.5%) | 224<br>(0.3%) | 7886<br>(2.7%) |
| 45 to 49 years | 5573<br>(11.7%) | 5014<br>(6.3%) | 2375<br>(3.0%) | 636<br>(0.8%) | 13598<br>(4.7%) |
| 50 to 54 years | 6455<br>(13.5%) | 6764<br>(8.4%) | 3660<br>(4.6%) | 1303<br>(1.6%) | 18182<br>(6.3%) |
| 55 to 59 years | 7192<br>(15.1%) | 8868<br>(11.1%) | 5679<br>(7.1%) | 2670<br>(3.3%) | 24409<br>(8.5%) |
| 60 to 64 years | 7006<br>(14.7%) | 10894<br>(13.6%) | 8388<br>(10.4%) | 5087<br>(6.4%) | 31375<br>(10.9%) |
| 65 to 69 years | 6824<br>(14.3%) | 13506<br>(16.8%) | 12589<br>(15.7%) | 9110<br>(11.4%) | 42029<br>(14.6%) |
| 70 to 74 years | 4898<br>(10.3%) | 11894<br>(14.8%) | 13808<br>(17.2%) | 12988<br>(16.2%) | 43588<br>(15.1%) |
| 75 to 79 years | 3092<br>(6.5%) | 9157<br>(11.4%) | 13026<br>(16.2%) | 15286<br>(19.1%) | 40561<br>(14.1%) |
| 80 to 84 years | 1829<br>(3.8%) | 6176<br>(7.7%) | 10245<br>(12.7%) | 15136<br>(18.9%) | 33386<br>(11.6%) |
| 85 to 89 years | 868<br>(1.8%) | 3411<br>(4.3%) | 6238<br>(7.8%) | 11071<br>(13.9%) | 21588<br>(7.5%) |
| 90 years and older | 335<br>(0.7%) | 1663<br>(2.1%) | 3246<br>(4.0%) | 6451<br>(8.1%) | 11695<br>(4.1%) |
| Gender (Female) | 23562<br>(49.4%) | 37167<br>(46.3%) | 37289<br>(46.4%) | 35665<br>(44.6%) | 133683<br>(46.4%) |
| *IMD* |  |  |  |  |  |
| 1 (least deprived) | 12,528<br>(26.3%) | 21563<br>(26.9%) | 17791<br>(22.1%) | 14294<br>(17.9%) | 66176<br>(23.0%) |
| 2 | 10824<br>(22.7%) | 18435<br>(23.0%) | 17247<br>(21.5%) | 15274<br>(19.1%) | 61780<br>(21.4%) |
| 3 | 9599<br>(20.2%) | 15708<br>(19.6%) | 15646<br>(19.5%) | 15445<br>(19.3%) | 56389<br>(19.6%) |
| 4 | 8262<br>(17.3%) | 13588<br>(16.9%) | 15194<br>(18.9%) | 16367<br>(20.5%) | 53411<br>(18.5%) |
| 5 (most deprived) | 6427<br>(13.5%) | 10908<br>(13.6%) | 14546<br>(18.1%) | 18560<br>(23.2%) | 50441<br>(17.5%) |
| *Ethnicity* |  |  |  |  |  |
| White | 43244 | 74056 | 74422 | 75107 | 266829 |
|  | (90.7%) | (92.3%) | (92.5%) | (93.9%) | (92.6%) |
| Asian | 1377 | 1980 | 2380 | 2225 | 7962 |
|  | (2.9%) | (2.5%) | (3.0%) | (2.8%) | (2.8%) |
| Black | 1128 | 2176 | 2142 | 1662 | 7108 |
|  | (2.4%) | (2.7%) | (2.7%) | (2.1%) | (2.5%) |
| Other | 414 | 509 | 430 | 331 | 1684 |
|  | (0.9%) | (0.6%) | (0.5%) | (0.4%) | (0.6%) |
| Mixed | 284 | 460 | 434 | 331 | 1509 |
|  | (0.6%) | (0.6%) | (0.5%) | (0.4%) | (0.5%) |
| Unknown | 1211 | 1046 | 642 | 306 | 3205 |
|  | (2.5%) | (1.3%) | (0.8%) | (0.4%) | (1.1%) |
| *Cancer type* |  |  |  |  |  |
| Bladder | 976 | 1934 | 2332 | 2782 | 8024 |
|  | (2.1%) | (2.4%) | (2.9%) | (3.5%) | (2.8%) |
| Brain | 861 | 1334 | 1067 | 763 | 4025 |
|  | (1.8%) | (1.7%) | (1.3%) | (1.0%) | (1.4%) |
| Breast | 7373 | 9697 | 7738 | 6350 | 31158 |
|  | (15.5%) | (12.1%) | (9.6%) | (7.9%) | (10.8%) |
| Cervix | 447 | 382 | 307 | 201 | 1337 |
|  | (0.9%) | (0.5%) | (0.4%) | (0.3%) | (0.5%) |
| Colon | 3289 | 5986 | 6409 | 6560 | 22244 |
|  | (6.9%) | (7.5%) | (8.0%) | (8.2%) | (7.7%) |
| HL | 275 | 315 | 283 | 207 | 1080 |
|  | (0.6%) | (0.4%) | (0.4%) | (0.3%) | (0.4%) |
| Larynx | 266 | 464 | 522 | 565 | 1817 |
|  | (0.6%) | (0.6%) | (0.7%) | (0.7%) | (0.6%) |
| Leukaemia | 1291 | 1967 | 2068 | 1990 | 7316 |
|  | (2.7%) | (2.5%) | (2.6%) | (2.5%) | (2.5%) |
| Liver | 463 | 886 | 1627 | 2207 | 5183 |
|  | (1.0%) | (1.1%) | (2.0%) | (2.8%) | (1.8%) |
| Lung | 4189 | 6926 | 10496 | 16578 | 38189 |
|  | (8.8%) | (8.6%) | (13.1%) | (20.7%) | (13.3%) |
| Melanoma | 2986 | 3876 | 2800 | 1901 | 11563 |
|  | (6.3%) | (4.8%) | (3.5%) | (2.4%) | (4.0%) |
| Mesothelioma | 304 | 663 | 693 | 671 | 2331 |
|  | (0.6%) | (0.8%) | (0.9%) | (0.8%) | (0.8%) |
| Myeloma | 781 | 1469 | 1471 | 1379 | 5100 |
|  | (1.6%) | (1.8%) | (1.8%) | (1.7%) | (1.8%) |
| NHL | 2016 | 3289 | 3356 | 2791 | 11452 |
|  | (4.2%) | (4.1%) | (4.2%) | (3.5%) | (4.0%) |
| Oesophagus | 1156 | 2097 | 2326 | 2498 | 8077 |
|  | (2.4%) | (2.6%) | (2.9%) | (3.1%) | (2.8%) |
| Oral | 1529 | 2126 | 2032 | 1775 | 7462 |
|  | (3.2%) | (2.7%) | (2.5%) | (2.2%) | (2.6%) |
| Other | 2871 | 4677 | 5522 | 6353 | 19423 |
|  | (6.0%) | (5.8%) | (6.9%) | (8.0%) | (6.7%) |
| Ovary | 1323 | 2087 | 1718 | 1242 | 6370 |
|  | (2.8%) | (2.6%) | (2.1%) | (1.6%) | (2.2%) |
| Pancreas | 1138 | 2143 | 2728 | 2893 | 8902 |
|  | (2.4%) | (2.7%) | (3.4%) | (3.6%) | (3.1%) |
| Prostate | 6718<br>(14.1%) | 15812<br>(19.7%) | 13535<br>(16.8%) | 10566<br>(13.2%) | 46631<br>(16.2%) |
| Rectum | 2185<br>(4.6%) | 3305<br>(4.1%) | 3206<br>(4.0%) | 2694<br>(3.4%) | 11390<br>(4.0%) |
| Renal | 1260<br>(2.6%) | 2534<br>(3.2%) | 2643<br>(3.3%) | 2653<br>(3.3%) | 9090<br>(3.2%) |
| Stomach | 738<br>(1.6%) | 1372<br>(1.7%) | 1615<br>(2.0%) | 1892<br>(2.4%) | 5617<br>(2.0%) |
| Testis | 425<br>(0.9%) | 318<br>(0.4%) | 169<br>(0.2%) | 52<br>(0.1%) | 964<br>(0.3%) |
| Thyroid | 586<br>(1.2%) | 936<br>(1.2%) | 619<br>(0.8%) | 309<br>(0.4%) | 2450<br>(0.9%) |
| Unspecified breast | 746<br>(1.6%) | 1003<br>(1.3%) | 628<br>(0.8%) | 383<br>(0.5%) | 2760<br>(1.0%) |
| Uterus | 1466<br>(3.1%) | 2629<br>(3.3%) | 2540<br>(3.2%) | 1707<br>(2.1%) | 8342<br>(2.9%) |

**Table 2.** Outcomes and routes to diagnosis for patients categorised in four morbidity burden groups.

|  | **No morbidity**<br>**burden (n=**<br>**47658)** | **Low morbidity**<br>**burden (n=**<br>**80227)** | **Medium**<br>**morbidity**<br>**burden (n=**<br>**80450)** | **High morbidity**<br>**burden (n=**<br>**79962)** | **Total (n=**<br>**288297)** |
| --- | --- | --- | --- | --- | --- |
|  | N (%) | N (%) | N (%) | N (%) | N (%) |
| *Outcomes* |  |  |  |  |  |
| 30-day mortality post-<br>diagnosis of cancer | 2273<br>(4.8%) | 4183<br>(5.2%) | 6662<br>(8.3%) | 11217<br>(14.0%) | 24335<br>(8.4%) |
| Urgent suspected cancer<br>referral route to diagnosis | 21883<br>(45.9%) | 35400<br>(44.1%) | 32855<br>(40.8%) | 27168<br>(34.0%) | 117306<br>(40.7%) |
| Emergency route to diagnosis | 7208<br>(15.1%) | 12246<br>(15.3%) | 16143<br>(20.1%) | 23052<br>(28.8%) | 58649<br>(20.3%) |
|  | n= 37844* | n = 63754* | n = 61818* | n = 57201* | n = 220617* |
| Advanced-stage cancer<br>diagnosis | 17950<br>(47.4%) | 29778<br>(46.7%) | 31526<br>(51.0%) | 31196<br>(54.5%) | 110450<br>(50.1%) |

\*276,956 patients had a recorded diagnosis of a stageable cancer in NCRAS. Information on stage was provided for 220,617 (79.7%) of these patients. The number of patients in each burden group is therefore lower than for other outcomes (Appendix A: Table A.2).

### Cancer stage

The main effects-only model showed that increasing morbidity burden was associated with reduced odds of advance-stage cancer (adjusted OR high vs. no morbidity burden: 0.72, 95% CI: 0.7 to 0.75, p<0.0001) (Figure 1; Appendix A: Table A.3). The interaction effect model suggested that for most cancers, increasing morbidity burden was associated with a decreasing likelihood of being diagnosed with advanced-stage cancer, while for some sites no significant associations were found (interaction p<0.0001) (Appendix A: Figure A.1).

### Emergency presentation route to diagnosis

A U-shaped relationship was seen such that patients with high morbidity burden were the most likely to be diagnosed via an emergency presentation in the main effects-only model, followed by patients with no morbidity burden (adjusted OR high vs. no morbidity burden: 1.19, 95% CI: 1.15 to 1.24, p<0.0001) (Figure 2; Appendix A: Table A.4). The interaction effects model showed varied associations for different cancers (Appendix A: Figure A.2). Patients with high morbidity burden were more likely to be diagnosed via an emergency presentation if they were diagnosed with non-Hodgkin lymphoma, melanoma, mesothelioma, prostate oral, ovarian, lung, pancreatic, stomach and oesophageal cancer (interaction p<0.0001).

**Figure. 2.**
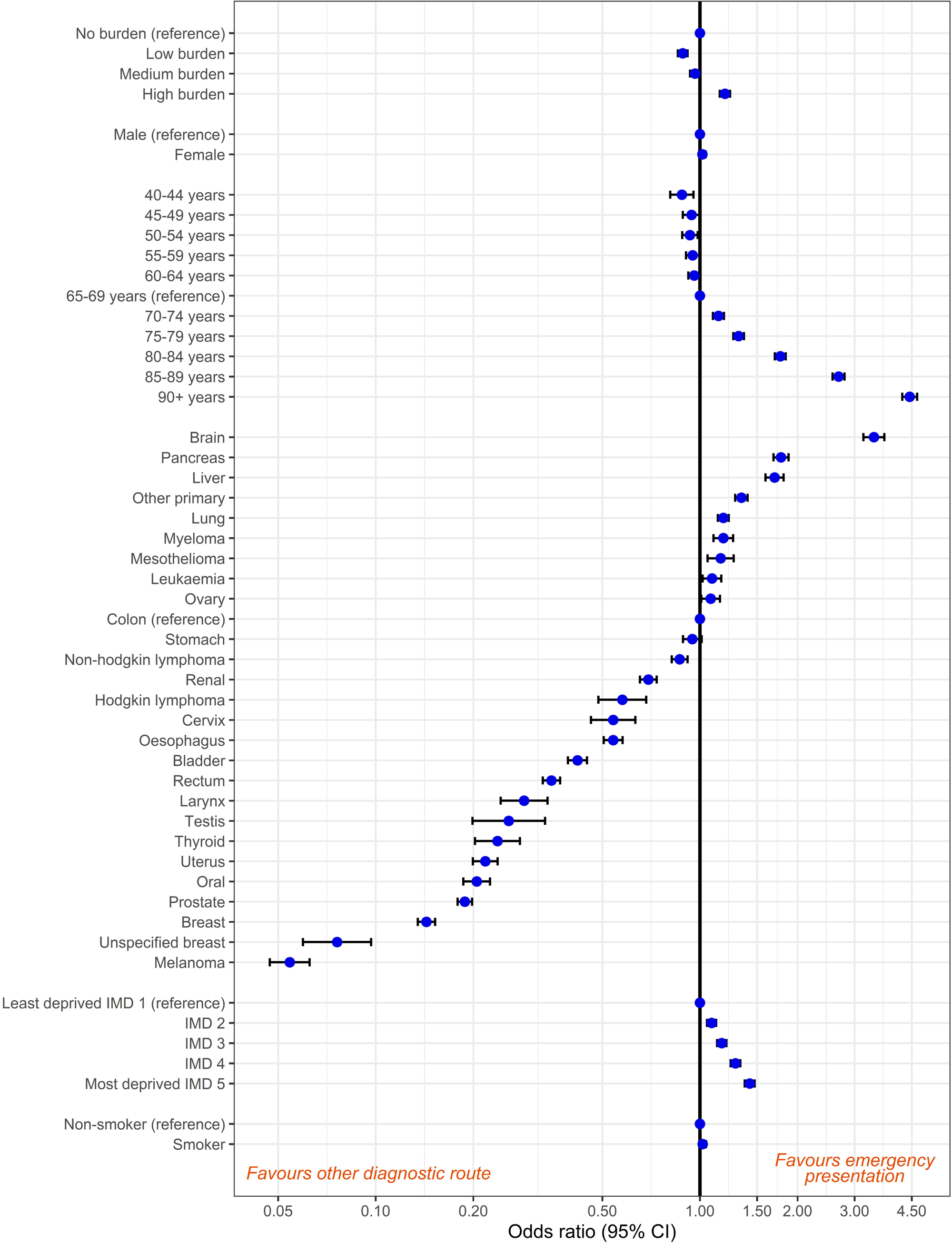
Associations between emergency route to diagnosis, patient characteristics, cancer site and morbidity burden*. *Confidence intervals were provided for all associations. However, narrow intervals may not be visible as they overlap with the dot indicating the odd ratio point estimate

### Urgent suspected cancer referral route to diagnosis

The main effects-only model showed that patients with higher morbidity burden were less likely to be diagnosed via an urgent suspected cancer referral (adjusted OR high vs. no morbidity burden: 0.75, 95% CI: 0.73 to 0.77, p<0.0001) (Figure 3; Appendix A: Table A.4). The interaction effects model showed that for nearly all cancers, patients with higher morbidity burden were less likely to be diagnosed via an urgent suspected cancer referral (interaction p<0.0001). For breast cancer, the no morbidity burden group was the least likely to be diagnosed via an urgent suspected cancer referral, and there were a number of sites where there was no clear relationship (melanoma, mesothelioma, testis, larynx, oral, and brain cancer) (Appendix A: Figure A.3).

**Figure. 3.**
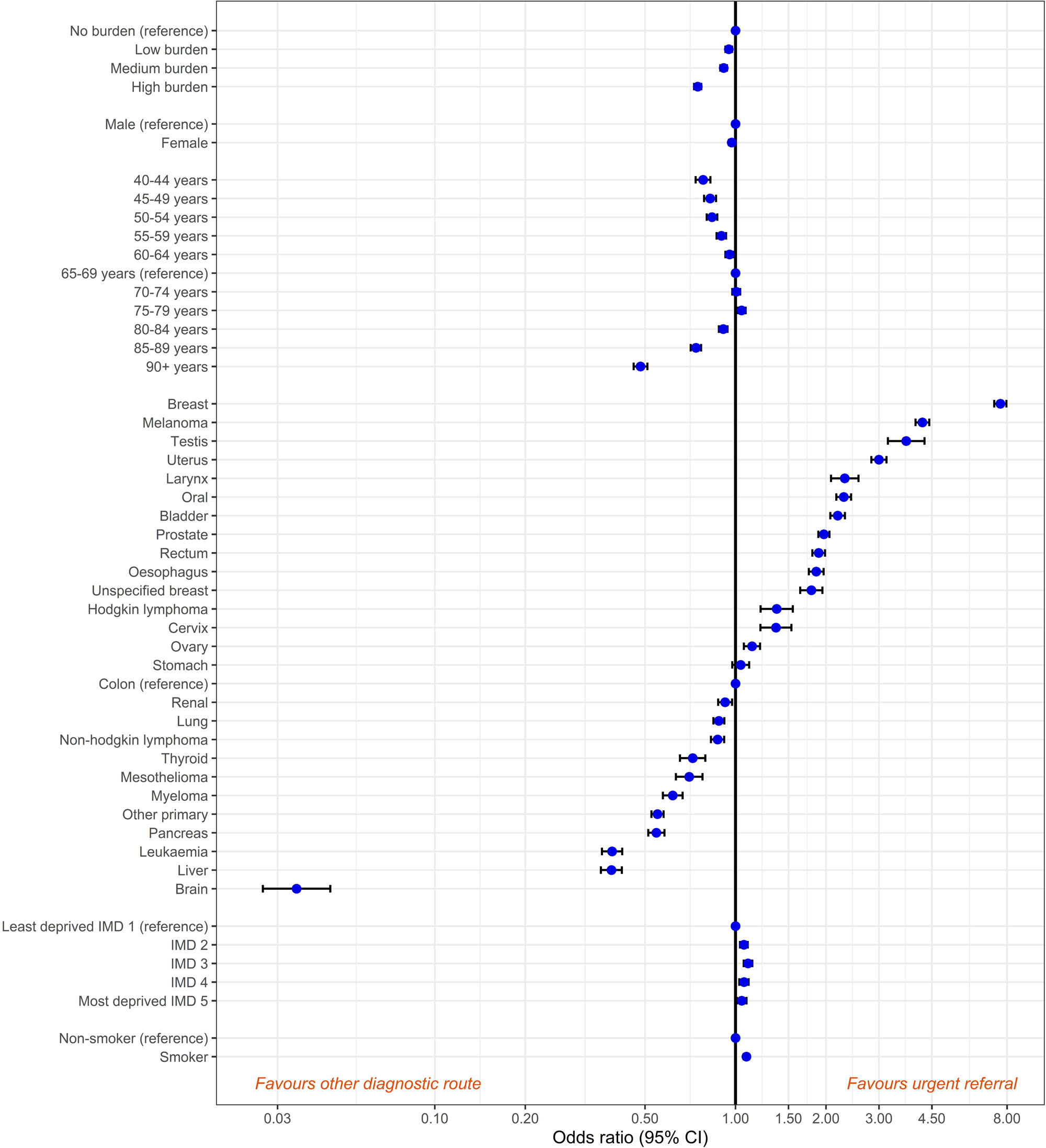
Associations between a suspected cancer referral route to diagnosis, patient characteristics, cancer site and morbidity burden*. *Confidence intervals were provided for all associations. However, narrow intervals may not be visible as they overlap with the dot indicating the odd ratio point estimate.

### 30-day mortality

A U-shaped relationship was seen such that patients with high morbidity burden were the most likely to die within 30 days after diagnosis (adjusted OR high vs. no morbidity burden: 1.20, 95% CI: 1.13 to 1.26, p<0.0001), followed by patients with no morbidity burden (Figure 4; Appendix A: Table A.3). The interaction effects model showed varied associations for different cancers. Patients with high morbidity burden were more likely to die within 30 days if they were diagnosed with leukaemia, ovarian cancer, non-Hodgkin lymphoma, prostate cancer, melanoma, cervix cancer, and brain cancer (interaction p<0.0001) (Appendix A: Figure A.4).

**Figure. 4.**
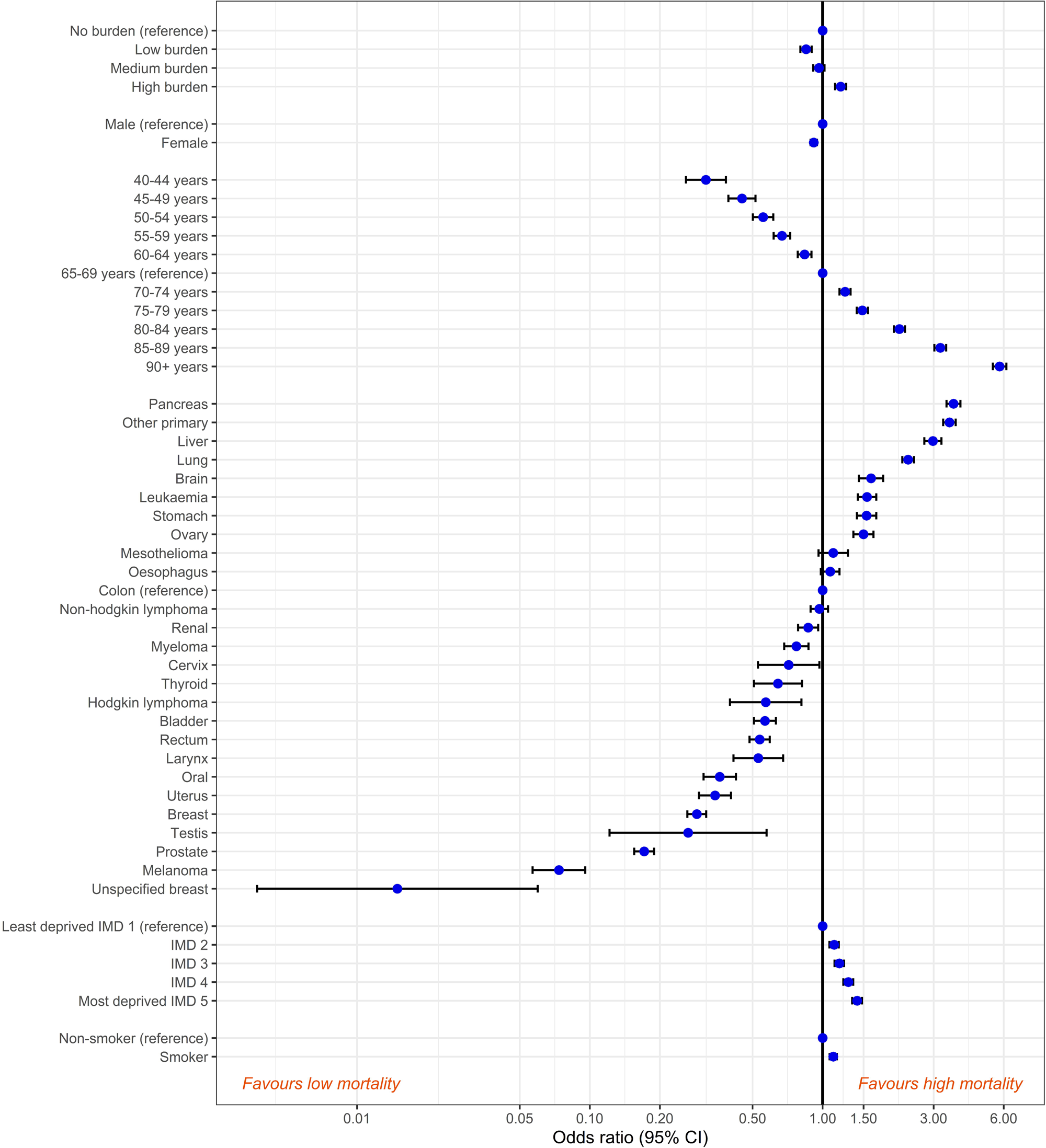
Associations between 30-day mortality, patient characteristics, cancer site and morbidity burden*. *Confidence intervals were provided for all associations. However, narrow intervals may not be visible as they overlap with the dot indicating the odd ratio point estimate.

### Sensitivity analyses

The main effects-only model using a composite outcome of advanced stage and emergency presentation suggested that patients with higher morbidity burden were less likely to be diagnosed with advanced stage as an emergency than patients without morbidity burden (p<0.0001) (Appendix A: Table A.5). Additional sensitivity analyses did not materially impact the findings (Appendix A).

## Discussion

### Statement of principal findings

This study is, to our knowledge, the largest and most comprehensive investigation into whether pre-existing morbidity is associated with cancer diagnostic outcomes and processes. Our study demonstrates that, contrary to expectations, increasing morbidity burden is associated with a reduction of advanced-stage cancer diagnoses, and that those without morbidities are the most likely to be diagnosed at an advanced stage. This applies to cancer in general, and is also seen across most cancer sites. Patients without morbidity burden were also more likely to be diagnosed as an emergency, and or to die within 30 days after diagnosis than patients with all but the highest morbidity burden. However, increasing morbidity burden was associated with a decreased proportion of cancers being diagnosed via urgent suspected cancer referrals, which was highest in the no-morbidity burden group.

### Comparison with existing literature and meaning of the study

A recent review showed associations between pre-existing morbidities and cancer stage in varying directions [13]. However, many of the included studies examined individual morbidity and cancer combinations [34–39]. Different mechanisms may be operating in different directions for specific morbidity and cancer combinations [13], which may not be visible when the burden of all morbidities is studied together as we have done here.

In line with previous research [13, 40–45], our findings also showed that patients with the highest morbidity burden were most likely to be diagnosed after an emergency presentation, and experience higher mortality. However, it has been less clearly reported that higher morbidity burden appears protective against advanced-stage diagnoses and that those without pre-existing morbidities are also commonly diagnosed as an emergency. One explanation for our findings is that patients with higher morbidity burden utilise health care more often than those without [46–49], receive more tests and investigations [19], and attend the emergency department more often [50], offering greater opportunities for diagnosing cancer early through a surveillance effect. This is unlike other mechanisms, where pre-existing morbidity may convey a disadvantage. Given emergency presentation is generally linked to worse patient outcomes, even when accounting for stage at diagnosis [28, 51], it is important to prioritise cancer diagnosis through non-emergency routes.

Finally, the present study suggested that patients with higher morbidity burden were less likely to be diagnosed via an urgent suspected cancer referral than patients with no morbidity burden. Previous evidence showed that patients with pre-existing morbidities diagnosed with colorectal cancer were less likely to be diagnosed after an urgent suspected cancer referral [52], and that patients with higher morbidity presenting with possible cancer features were less likely to receive an urgent suspected cancer referral [53]. It is possible that GPs are less likely to recognise that patients with morbidity need to be referred. Alternatively, regular monitoring of morbidities may result in symptoms being investigated before they qualify for an urgent suspected cancer referral, or they are investigated as a result of morbidity management in secondary care.

### Strengths and limitations

We used a large, validated, longitudinal dataset of patient health records [22, 23], which allowed us to include a large number of cancer patients across all cancer sites. Complete patient records meant that we were able to include a broad range of morbidities, and compare several morbidity measures.

There are, however, some limitations. Morbidity was identified using medical or prescription codes in patients’ health records. Some morbidities may have been missed because they were not recorded or were recorded in ‘free/uncoded text’ (inaccessible to researchers). CPRD studies suggest, however, that there is a high overlap between free-text and diagnoses codes [54]. Although our morbidity burden measure, the CMS, has been validated, it is limited to 37 common morbidities [29]. Patients may have had other or additional morbidities. Sensitivity analyses using an alternative morbidity measure (the ACG system) produced very similar results, reducing such concerns. None of these measures, however, take into account morbidity type, severity, or control, which could affect the cancer diagnostic process. Future research is needed to investigate how specific pre-existing morbidities, or the severity of these morbidities affect outcomes. Finally, morbidity was measured a year before diagnosis to exclude potential misdiagnosis of cancer symptoms as a new morbidity. This study is therefore unable to give insight into how newly diagnosed morbidities and their management could affect the cancer diagnostic process.

## Conclusions

Contrary to expectations, not having pre-existing morbidities was associated with increased risk of advanced-stage diagnosis, emergency presentations and 30-day mortality. This may reflect heightened surveillance of patients with morbidity being protective against later advanced-stage cancer diagnoses. Although patients with high morbidity burden were more likely to be diagnosed after an emergency presentation or to die within 30 days of diagnosis, increasing morbidity burden was protective against advanced stage at diagnosis. This indicates that poorer outcomes in those with pre-existing morbidity may be caused by the pre-existing morbidities, rather than a more advanced stage diagnosis, emphasising the importance of comprehensive care for individuals with multimorbidity.

## Statements & Declarations

### Author Contributions

Conceptualization, Luke Mounce, Jose Valderas and Gary Abel; Data curation, Bianca Wiering; Formal analysis, Bianca Wiering; Funding acquisition, Luke Mounce, Jose Valderas and Gary Abel; Investigation, Bianca Wiering and David Shotter; Methodology, Bianca Wiering, Luke Mounce, Sarah Price, David Shotter and Gary Abel; Project administration, Luke Mounce and Gary Abel; Resources, Sarah Price, David Shotter, Jose Valderas, Samuel Merriel, Sarah Moore, Rupert Payne, Cristina Renzi, Georgios Lyratzopoulos and Willie Hamilton; Software, Bianca Wiering and David Shotter; Supervision, Luke Mounce and Gary Abel; Validation, Luke Mounce, Sarah Price and Gary Abel; Visualization, Bianca Wiering, Luke Mounce, Rupert Payne, Georgios Lyratzopoulos, Willie Hamilton and Gary Abel; Writing – original draft, Bianca Wiering; Writing – review & editing, Bianca Wiering, Luke Mounce, Sarah Price, David Shotter, Jose Valderas, Samuel Merriel, Sarah Moore, Leon Farmer, Christian Von Wagner, Rupert Payne, Cristina Renzi, Georgios Lyratzopoulos, Willie Hamilton and Gary Abel.

### Ethics approval

The National Research Ethics Service (NRES) has granted ethical approval for observational research using anonymised CPRD data. Observational studies using anonymised CPRD data are therefore not required to obtain study-specific ethical approval. Study protocol approval was granted by the CPRD Independent Scientific Advisory Committee (protocol 20_126R).

### Consent to participate

Patient consent was waived due to the nature of the data collection. GP practices choose to share patient data with CPRD for public health research purposes. No information that can identify a patient is ever sent to CPRD from the contributing GP practices. Because patients can’t be identified from data a GP practice sends to CPRD, the GP practice does not need to seek a patient’s consent to share data with CPRD. Individual patients can opt-out of sharing their data for research. CPRD does not collect data for these patients.

### Consent to publish

No identifiable patient data was made available to the research team by CPRD. Consent for publication has therefore not been sought.

### Data Availability Statement

Restrictions apply to the availability of these data. Routinely collected patient electronic health data was provided by CPRD. Access to data from CPRD is subject to a licence agreement and protocol approval.

## Acknowledgments

This study is based on data from the Clinical Practice Research Datalink obtained under licence from the UK Medicines and Healthcare products Regulatory Agency. The data is provided by patients and collected by the NHS as part of their care and support. The interpretation and conclusions contained in this study are those of the author/s alone. The study is funded by the NIHR Programme Grants for Applied Research (PGfAR) SPOtting Cancer among Comorbidities (SPOCC) programme: supporting clinical decision making in patients with symptoms of cancer and pre-existing conditions (NIHR201070). The views expressed are those of the author(s) and not necessarily those of the NIHR or the Department of Health and Social Care. GL was supported by a Cancer Research UK Fellowship C18081/A18180. SWDM is supported by the NIHR Manchester Biomedical Research Centre (NIHR203308). SM is supported by a doctoral fellowship for primary care clinicians awarded by Wellcome, grant number PMHG1A4. Additionally, we would like to thank Johns Hopkins University for providing the ACG software.

## Declaration of generative AI in scientific writing

generative AI and AI-assisted technologies were not used in the preparation of this work.

## Abbreviations

The following abbreviations are used in this manuscript:

CPRD: Clinical Practice Research Datalink
NCRAS: National Cancer Registration and Analysis Service
HES: Hospital Episode Statistics
IMD: Index of Multiple Deprivation
ONS: Office for National Statistics
NRES: National Research Ethics Service
NIHR: National Institute for Health and Care Research
PPI: Patient and Public Involvement
SPOCC: SPOtting Cancer among Comorbidities
TNM: Tumour, Node, Metastasis
CMS: Cambridge Multimorbidity Score
GP: General Practitioner
ACG: Adjusted Clinical Groups

## Supplementary Materials

The following supporting information can be found in Appendix A: Table A.1: ICD-10 codes for each cancer site; Table A.2: Missing cancer stage for stageable cancers by morbidity burden group; Table A.3: Main effect only model associations between cancer stage, 30-day mortality, patient characteristics, and cancer type (CMS morbidity burden); Table A.4: Main effect only model associations between emergency presentation route to diagnosis, urgent (two-week-wait) referral route to diagnosis, patient characteristics, and cancer type (CMS morbidity burden); Table A.5: Sensitivity analysis: Associations between a composite outcome of advanced cancer stage and emergency presentation route to diagnosis and patient characteristics (CMS morbidity burden); Table A.6: Sensitivity analysis: Associations between 30-day mortality, emergency presentation route to diagnosis, urgent (two-week-wait) referral route to diagnosis and morbidity burden level with cancer stage included in the model (CMS morbidity burden); Table A.7: Sensitivity analysis: Associations between cancer stage, 30-day mortality and patient characteristics (ACG System); Table A.8: Sensitivity analysis: Associations between emergency presentation route to diagnosis, urgent (two-week-wait) referral route to diagnosis, and patient characteristics (ACG System); Table A.9: Sensitivity analysis: Associations between cancer stage, 30-day mortality, and patient characteristics (morbidity count based on CMS); Table A.10: Sensitivity analysis: Associations between emergency presentation route to diagnosis, urgent (two-week-wait) referral route to diagnosis, and patient characteristics (morbidity count based on CMS); Figure A.1: Associations between cancer stage and morbidity burden by cancer type; Figure A.2: Associations between emergency route to diagnosis and morbidity burden by cancer type; Figure A.3: Associations between an urgent suspected cancer referral route to diagnosis and morbidity burden by cancer type; Figure A.4: Associations between 30-day mortality and morbidity burden by cancer type

