## Appendix A for "The impact of morbidity burden on cancer diagnosis; a retrospective cohort study in England"

**Table A.1. ICD-10 codes for each cancer site**

| Cancer site | ICD-10 code |
|---|---|
| Lip, oral cavity and pharynx | C00, C01, C02, C03, C04, C05, C06, C07, C08, C09, C10, C11, C12, C13, C14 |
| Oesophagus | C15 |
| Liver | C22 |
| Stomach | C16 |
| Colon | C18 |
| Rectum | C19, C20 |
| Pancreas | C25 |
| Larynx | C32 |
| Lung | C33, C34 |
| Melanoma | C43 |
| Mesothelioma | C45 |
| Breast | C50 |
| Cervix | C53 |
| Uterus | C54, C55 |
| Ovary | C56, C57 |
| Prostate | C61 |
| Testis | C62 |
| Kidney | C64 |
| Bladder | C67 |
| Brain | C71 |
| Thyroid | C73 |
| Hodgkin lymphoma | C81 |
| Non-Hodgkin lymphoma | C82, C83, C84, C85 |
| Myeloma | C90 |
| Leukaemia | C91, C92, C93, C94, C95 |
| Unspecified breast | D05 |

**Table A.2. Missing cancer stage for stageable cancers by morbidity burden group**

| | Stageable cancers | Missing cancer stage |
|---|---|---|
| | N (%) | N (%) |
| No morbidity burden | 45506 (16.4%) | 7662 (16.8%) |
| Low morbidity burden | 76926 (27.8%) | 13172 (17.1%) |
| Medium morbidity burden | 77315 (27.9%) | 15497 (20%) |
| High morbidity burden | 77209 (27.9%) | 20008 (25.9%) |
| Total | 276956 (100%) | 56339 (20.3%) |

**Table A.3. Main effect only model associations between cancer stage, 30-day mortality, patient characteristics, and cancer type (CMS morbidity burden)**

| | Advanced-stage cancer diagnosis | | | | 30-day mortality | | | |
|---|---|---|---|---|---|---|---|---|
| | Odds ratio | Lower 95% confidence interval | Upper 95% confidence interval | P | Odds ratio | Lower 95% confidence interval | Upper 95% confidence interval | P |
| Bladder | 0.24 | 0.23 | 0.26 | P<0.0001 | 0.57 | 0.51 | 0.63 | P<0.0001 |
| Brain | | | | | 1.61 | 1.43 | 1.82 | |
| Breast | 0.21 | 0.20 | 0.22 | | 0.29 | 0.26 | 0.32 | |
| Cervix | 0.45 | 0.39 | 0.51 | | 0.72 | 0.53 | 0.97 | |
| Colon | 1.00 | 1.00 | 1.00 | | 1.00 | 1.00 | 1.00 | |
| HL | 1.03 | 0.90 | 1.18 | | 0.57 | 0.40 | 0.81 | |
| Larynx | 0.70 | 0.62 | 0.78 | | 0.53 | 0.41 | 0.68 | |
| Leukaemia | | | | | 1.55 | 1.42 | 1.70 | |
| Liver | 1.68 | 1.53 | 1.85 | | 2.98 | 2.74 | 3.24 | |
| Lung | 2.32 | 2.22 | 2.41 | | 2.33 | 2.20 | 2.47 | |
| Melanoma | 0.08 | 0.08 | 0.09 | | 0.07 | 0.06 | 0.10 | |
| Mesothelioma | 1.66 | 1.45 | 1.92 | | 1.11 | 0.96 | 1.28 | |
| Myeloma | 0.36 | 0.32 | 0.41 | | 0.77 | 0.68 | 0.87 | |
| NHL | 1.61 | 1.52 | 1.70 | | 0.97 | 0.89 | 1.05 | |
| Oesophagus | 2.18 | 2.04 | 2.33 | | 1.08 | 0.98 | 1.18 | |
| Oral | 2.07 | 1.93 | 2.21 | | 0.36 | 0.31 | 0.42 | |
| Other | 1.14 | 1.07 | 1.21 | | 3.51 | 3.30 | 3.73 | |
| Ovary | 1.31 | 1.23 | 1.40 | | 1.50 | 1.36 | 1.66 | |
| Pancreas | 2.78 | 2.60 | 2.97 | | 3.65 | 3.41 | 3.91 | |
| Prostate | 0.54 | 0.52 | 0.56 | | 0.17 | 0.16 | 0.19 | |
| Rectum | 1.00 | 0.95 | 1.05 | | 0.54 | 0.49 | 0.59 | |
| Renal | 0.66 | 0.62 | 0.70 | | 0.87 | 0.79 | 0.96 | |
| Stomach | 1.82 | 1.69 | 1.97 | | 1.54 | 1.40 | 1.70 | |
| Testis | 0.07 | 0.06 | 0.10 | | 0.26 | 0.12 | 0.58 | |
| Thyroid | 0.54 | 0.48 | 0.60 | | 0.64 | 0.51 | 0.82 | |
| Unspecified breast | 1.00 | 1.00 | 1.00 | | 0.01 | 0.00 | 0.06 | |
| Uterus | 0.20 | 0.19 | 0.21 | | 0.35 | 0.29 | 0.40 | |
| *Morbidity burden* | | | | | | | | |
| No morbidity burden | 1.00 | 1.00 | 1.00 | p<0.0001 | 1.00 | 1.00 | 1.00 | p<0.0001 |
| Low morbidity burden | 0.83 | 0.81 | 0.86 | | 0.85 | 0.80 | 0.90 | |
| Medium morbidity burden | 0.81 | 0.78 | 0.83 | | 0.96 | 0.91 | 1.02 | |
| High morbidity burden | 0.72 | 0.70 | 0.75 | | 1.20 | 1.13 | 1.26 | |
| Gender (Female) | 0.88 | 0.85 | 0.90 | p<0.0001 | 0.92 | 0.89 | 0.95 | p<0.0001 |
| *Age* | | | | | | | | |
| 40 to 44 years | 0.65 | 0.61 | 0.70 | p<0.0001 | 0.32 | 0.26 | 0.38 | p<0.0001 |
| 45 to 49 years | 0.78 | 0.74 | 0.82 | | 0.45 | 0.39 | 0.52 | |
| 50 to 54 years | 0.85 | 0.81 | 0.89 | | 0.56 | 0.50 | 0.61 | |
| 55 to 59 years | 0.88 | 0.85 | 0.92 | | 0.67 | 0.62 | 0.73 | |
| 60 to 64 years | 0.95 | 0.91 | 0.98 | | 0.84 | 0.78 | 0.90 | |
| 65 to 69 years | 1.00 | 1.00 | 1.00 | | 1.00 | 1.00 | 1.00 | |
| 70 to 74 years | 1.04 | 1.01 | 1.07 | | 1.25 | 1.18 | 1.32 | |
| 75 to 79 years | 1.10 | 1.06 | 1.14 | | 1.48 | 1.40 | 1.57 | |
| 80 to 84 years | 1.20 | 1.15 | 1.25 | | 2.14 | 2.02 | 2.26 | |
| 85 to 89 years | 1.37 | 1.31 | 1.43 | | 3.20 | 3.02 | 3.40 | |
| 90 years and older | 1.51 | 1.41 | 1.61 | | 5.76 | 5.39 | 6.16 | |
| *IMD* | | | | | | | | |
| 1 (least deprived | 1.00 | 1.00 | 1.00 | p<0.0001 | 1.00 | 1.00 | 1.00 | p<0.0001 |
| 2 | 1.02 | 0.99 | 1.05 | | 1.12 | 1.07 | 1.17 | |
| 3 | 1.04 | 1.01 | 1.08 | | 1.18 | 1.13 | 1.24 | |
| 4 | 1.09 | 1.06 | 1.13 | | 1.29 | 1.23 | 1.35 | |
| 5 (most deprived) | 1.12 | 1.09 | 1.16 | | 1.41 | 1.34 | 1.48 | |
| Smoking history | 1.05 | 1.03 | 1.08 | p<0.0001 | 1.11 | 1.08 | 1.16 | p<0.0001 |

**Table A.4. Main effect only model associations between emergency presentation route to diagnosis, urgent (two-week-wait) referral route to diagnosis, patient characteristics, and cancer type (CMS morbidity burden)**

| | **Emergency presentation** | | | | **Urgent suspected cancer referral** | | | |
|---|---|---|---|---|---|---|---|---|
| | Odds ratio | Lower 95% confidence interval | Upper 95% confidence interval | P | Odds ratio | Lower 95% confidence interval | Upper 95% confidence interval | P |
| Bladder | 0.42 | 0.39 | 0.45 | P<0.0001 | 2.19 | 2.07 | 2.31 | P<0.0001 |
| Brain | 3.44 | 3.19 | 3.71 | | 0.03 | 0.03 | 0.04 | |
| Breast | 0.14 | 0.13 | 0.15 | | 7.60 | 7.25 | 7.96 | |
| Cervix | 0.54 | 0.46 | 0.63 | | 1.36 | 1.21 | 1.54 | |
| Colon | 1.00 | 1.00 | 1.00 | | 1.00 | 1.00 | 1.00 | |
| HL | 0.58 | 0.49 | 0.68 | | 1.37 | 1.21 | 1.55 | |
| Larynx | 0.29 | 0.24 | 0.34 | | 2.31 | 2.08 | 2.56 | |
| Leukaemia | 1.09 | 1.02 | 1.16 | | 0.39 | 0.36 | 0.42 | |
| Liver | 1.70 | 1.59 | 1.81 | | 0.39 | 0.36 | 0.42 | |
| Lung | 1.18 | 1.14 | 1.23 | | 0.88 | 0.84 | 0.92 | |
| Melanoma | 0.05 | 0.05 | 0.06 | | 4.18 | 3.97 | 4.41 | |
| Mesothelioma | 1.16 | 1.06 | 1.27 | | 0.70 | 0.63 | 0.78 | |
| Myeloma | 1.18 | 1.10 | 1.27 | | 0.62 | 0.57 | 0.67 | |
| NHL | 0.87 | 0.82 | 0.92 | | 0.87 | 0.83 | 0.92 | |
| Oesophagus | 0.54 | 0.51 | 0.58 | | 1.86 | 1.75 | 1.96 | |
| Oral | 0.20 | 0.19 | 0.23 | | 2.29 | 2.16 | 2.43 | |
| Other | 1.34 | 1.28 | 1.40 | | 0.55 | 0.53 | 0.58 | |
| Ovary | 1.08 | 1.01 | 1.15 | | 1.13 | 1.07 | 1.21 | |
| Pancreas | 1.78 | 1.69 | 1.88 | | 0.55 | 0.51 | 0.58 | |
| Prostate | 0.19 | 0.18 | 0.20 | | 1.97 | 1.89 | 2.05 | |
| Rectum | 0.35 | 0.33 | 0.37 | | 1.89 | 1.80 | 1.99 | |
| Renal | 0.69 | 0.65 | 0.74 | | 0.92 | 0.88 | 0.97 | |
| Stomach | 0.95 | 0.89 | 1.01 | | 1.04 | 0.98 | 1.11 | |
| Testis | 0.26 | 0.20 | 0.33 | | 3.69 | 3.21 | 4.25 | |
| Thyroid | 0.24 | 0.20 | 0.28 | | 0.72 | 0.65 | 0.79 | |
| Unspecified breast | 0.08 | 0.06 | 0.10 | | 1.79 | 1.64 | 1.95 | |
| Uterus | 0.22 | 0.20 | 0.24 | | 3.00 | 2.83 | 3.18 | |
| *Morbidity burden* | | | | | | | | |
| No morbidity burden | 1.00 | 1.00 | 1.00 | p<0.0001 | 1.00 | 1.00 | 1.00 | p<0.0001 |
| Low morbidity burden | 0.89 | 0.86 | 0.92 | | 0.95 | 0.93 | 0.98 | |
| Medium morbidity burden | 0.97 | 0.93 | 1.00 | | 0.91 | 0.89 | 0.94 | |
| High morbidity burden | 1.19 | 1.15 | 1.24 | | 0.75 | 0.73 | 0.77 | |
| Gender (Female) | 1.02 | 0.99 | 1.04 | 0.1397 | 0.97 | 0.95 | 0.99 | 0.0104 |
| *Age* | | | | | | | | |
| 40 to 44 years | 0.88 | 0.81 | 0.96 | p<0.0001 | 0.78 | 0.74 | 0.83 | p<0.0001 |
| 45 to 49 years | 0.94 | 0.89 | 1.00 | | 0.82 | 0.79 | 0.86 | |
| 50 to 54 years | 0.93 | 0.88 | 0.98 | | 0.84 | 0.80 | 0.87 | |
| 55 to 59 years | 0.95 | 0.91 | 1.00 | | 0.90 | 0.87 | 0.93 | |
| 60 to 64 years | 0.96 | 0.92 | 1.00 | | 0.96 | 0.93 | 0.99 | |
| 65 to 69 years | 1.00 | 1.00 | 1.00 | | 1.00 | 1.00 | 1.00 | |
| 70 to 74 years | 1.14 | 1.10 | 1.19 | | 1.01 | 0.98 | 1.04 | |
| 75 to 79 years | 1.32 | 1.27 | 1.37 | | 1.05 | 1.01 | 1.08 | |
| 80 to 84 years | 1.77 | 1.70 | 1.84 | | 0.91 | 0.88 | 0.94 | |
| 85 to 89 years | 2.68 | 2.57 | 2.79 | | 0.74 | 0.71 | 0.77 | |
| 90 years and older | 4.43 | 4.21 | 4.67 | | 0.48 | 0.46 | 0.51 | |
| *IMD* | | | | | | | | |
| 1 (least deprived | 1.00 | 1.00 | 1.00 | p<0.0001 | 1.00 | 1.00 | 1.00 | p<0.0001 |
| 2 | 1.09 | 1.05 | 1.12 | | 1.07 | 1.04 | 1.10 | |
| 3 | 1.17 | 1.13 | 1.21 | | 1.10 | 1.06 | 1.14 | |
| 4 | 1.29 | 1.24 | 1.33 | | 1.07 | 1.03 | 1.11 | |
| 5 (most deprived) | 1.42 | 1.38 | 1.48 | | 1.05 | 1.01 | 1.09 | |

| | | | | | | | | |
|---|---|---|---|---|---|---|---|---|
| Smoking history | 1.02 | 1.00 | 1.05 | 0.1083 | 1.09 | 1.06 | 1.11 | p<0.0001 |

### Sensitivity analyses

#### Methods

A composite outcome was created which combined advanced stage with emergency route to diagnosis, with a value of one for patients with both advanced-stage cancer and an emergency route to diagnosis, and zero for all other patients. A main effects-only model was run as above with the composite outcome. Furthermore, as morbidity burden's association with 30-day mortality and routes to diagnosis may be affected by cancer stage, main effects-only models for these outcomes were rerun with cancer stage included as a covariate. Finally, besides CMS, two alternative morbidity measures were used to corroborate results. The first was a patient-level count of morbidities included in the CMS, as a condition count is a commonly used morbidity measure (32), and the second was the Johns Hopkins Adjusted Clinical Groups (ACG) system. The ACG system is population health analytics software developed for the United States. It uses primary and secondary care data to establish health care utilisation and predict future risk (33), and performs well compared to morbidity measures (34). CPRD and HES data was inputted and from the output we used the variable resource utilisation band. This variable had six bands (bands 0-5) with the lowest band indicating low resource utilisation, and the highest high resource utilisation. All analyses were conducted using the statistical software Stata v.17 (35), while figures 1 to 4 were produced using R (36).

#### Results

The main effects-only model using a composite outcome of advanced stage and emergency presentation suggested that patients with higher morbidity burden were less likely to be diagnosed with both advanced stage and after an emergency presentation than patients without morbidity burden (p<0.0001) (Appendix A: Table A.5).

Main effects-only analyses for 30-day mortality and routes to diagnosis with cancer stage included in the model, showed that patients with high morbidity were more likely to be diagnosed after an emergency presentation or to die within 30 days of diagnosis, and were less likely to be diagnosed after an urgent suspected cancer referral than patients without morbidity burden (p<0.0001) (Appendix A: Table A.6).

Main effects-only models using the ACG resource utilisation score, or a condition count showed similar results to the models using the CMS, with patients with higher morbidity less likely to be diagnosed with advanced cancer, or after an urgent suspected cancer referral (p<0.0001). Patients with high morbidity burden were more likely to die within 30 days of being diagnosed or to be diagnosed after an emergency presentation (p<0.0001) (Appendix A: Table A.7-10).

**Table A.5. Sensitivity analysis: Associations between a composite outcome of advanced cancer stage and emergency presentation route to diagnosis and patient characteristics (CMS morbidity burden).***

**Advanced-stage cancer diagnosis and emergency presentation route to diagnosis**

| | Odds ratio | Lower 95% confidence interval | Upper 95% confidence interval | P |
|---|---|---|---|---|
| *Morbidity burden* | | | | |
| No morbidity burden | 1.00 | 1.00 | 1.00 | p<0.0001 |
| Low morbidity burden | 0.87 | 0.83 | 0.91 | |
| Medium morbidity burden | 0.89 | 0.86 | 0.94 | |
| High morbidity burden | 0.93 | 0.88 | 0.97 | |
| Gender (Female) | 0.99 | 0.96 | 1.02 | 0.4550 |
| *Age* | | | | |
| 40 to 44 years | 0.85 | 0.76 | 0.95 | p<0.0001 |
| 45 to 49 years | 0.92 | 0.84 | 1.00 | |
| 50 to 54 years | 0.97 | 0.90 | 1.04 | |
| 55 to 59 years | 0.92 | 0.87 | 0.98 | |
| 60 to 64 years | 0.95 | 0.90 | 1.00 | |
| 65 to 69 years | 1.00 | 1.00 | 1.00 | |
| 70 to 74 years | 1.12 | 1.06 | 1.17 | |
| 75 to 79 years | 1.17 | 1.11 | 1.23 | |
| 80 to 84 years | 1.40 | 1.33 | 1.47 | |
| 85 to 89 years | 1.61 | 1.53 | 1.71 | |
| 90 years and older | 1.77 | 1.65 | 1.89 | |
| *IMD* | | | | |
| 1 (least deprived | 1.00 | 1.00 | 1.00 | p<0.0001 |
| 2 | 1.07 | 1.02 | 1.12 | |
| 3 | 1.15 | 1.10 | 1.20 | |
| 4 | 1.27 | 1.21 | 1.32 | |
| 5 (most deprived) | 1.39 | 1.33 | 1.45 | |
| Smoking history | 1.01 | 0.97 | 1.04 | 0.7395 |

*Analyses were controlled for cancer site, patient age, gender, deprivation quintile, year of diagnosis, and history of smoking

**Table A.6. Sensitivity analysis: Associations between 30-day mortality, emergency presentation route to diagnosis, urgent (two-week-wait) referral route to diagnosis and morbidity burden level with cancer stage included in the model (CMS morbidity burden). ***

| | 30-day mortality | | | | Emergency presentation | | | |
|---|---|---|---|---|---|---|---|---|
| | Odds ratio | Lower 95% confidence interval | Upper 95% confidence interval | P | Odds ratio | Lower 95% confidence interval | Upper 95% confidence interval | P |
| *Morbidity burden* | | | | | | | | |
| No morbidity burden | 1.00 | 1.00 | 1.00 | P<0.0001 | 1.00 | 1.00 | 1.00 | P<0.0001 |
| Low morbidity burden | 0.89 | 0.82 | 0.96 | | 0.90 | 0.86 | 0.94 | |
| Medium morbidity burden | 1.00 | 0.92 | 1.08 | | 0.98 | 0.94 | 1.02 | |
| High morbidity burden | 1.25 | 1.16 | 1.35 | | 1.21 | 1.16 | 1.27 | |

| | Urgent suspected cancer referral | | | |
|---|---|---|---|---|
| | Odds ratio | Lower 95% confidence interval | Upper 95% confidence interval | P |
| *Morbidity burden* | | | | |
| No morbidity burden | 1.00 | 1.00 | 1.00 | P<0.0001 |
| Low morbidity burden | 0.95 | 0.92 | 0.98 | |
| Medium morbidity burden | 0.91 | 0.89 | 0.94 | |
| High morbidity burden | 0.77 | 0.75 | 0.79 | |

*Analyses were controlled for cancer stage, cancer site, patient age, gender, deprivation quintile, year of diagnosis, and history of smoking.

**Table A.7. Sensitivity analysis: Associations between cancer stage, 30-day mortality and patient characteristics (ACG System). ***

| | Advanced-stage cancer diagnosis | | | | 30-day mortality | | | |
|---|---|---|---|---|---|---|---|---|
| | Odds ratio | Lower 95% confidence interval | Upper 95% confidence interval | P | Odds ratio | Lower 95% confidence interval | Upper 95% confidence interval | P |
| *Morbidity* | | | | | | | | |
| Resource | | | | P<0.0001 | | | | P<0.0001 |
| utilisation band 0 | 1.00 | 1.00 | 1.00 | | 1.00 | 1.00 | 1.00 | |
| Resource utilisation band 1 | 0.91 | 0.84 | 0.98 | | 0.88 | 0.77 | 1.01 | |
| Resource utilisation band 2 | 0.81 | 0.75 | 0.87 | | 0.66 | 0.58 | 0.76 | |
| Resource utilisation band 3 | 0.71 | 0.65 | 0.76 | | 0.68 | 0.60 | 0.77 | |
| Resource utilisation band 4 | 0.62 | 0.57 | 0.67 | | 0.79 | 0.69 | 0.89 | |
| Resource utilisation band 5 | 0.61 | 0.56 | 0.66 | | 1.02 | 0.89 | 1.16 | |
| Gender (Female) | 0.88 | 0.86 | 0.90 | P<0.0001 | 0.92 | 0.89 | 0.95 | P<0.0001 |
| *Age* | | | | | | | | |
| 40 to 44 years | 0.65 | 0.61 | 0.70 | P<0.0001 | 0.29 | 0.24 | 0.36 | P<0.0001 |
| 45 to 49 years | 0.78 | 0.74 | 0.82 | | 0.42 | 0.37 | 0.48 | |
| 50 to 54 years | 0.85 | 0.82 | 0.89 | | 0.53 | 0.48 | 0.58 | |
| 55 to 59 years | 0.88 | 0.85 | 0.92 | | 0.64 | 0.59 | 0.70 | |
| 60 to 64 years | 0.95 | 0.92 | 0.98 | | 0.82 | 0.77 | 0.88 | |
| 65 to 69 years | 1.00 | 1.00 | 1.00 | | 1.00 | 1.00 | 1.00 | |
| 70 to 74 years | 1.04 | 1.01 | 1.07 | | 1.27 | 1.20 | 1.35 | |
| 75 to 79 years | 1.10 | 1.06 | 1.13 | | 1.53 | 1.45 | 1.62 | |
| 80 to 84 years | 1.20 | 1.15 | 1.24 | | 2.22 | 2.11 | 2.35 | |
| 85 to 89 years | 1.37 | 1.31 | 1.43 | | 3.34 | 3.15 | 3.54 | |
| 90 years and older | 1.50 | 1.41 | 1.61 | | 6.02 | 5.63 | 6.43 | |
| *IMD* | | | | | | | | |
| 1 (least deprived) | 1.00 | 1.00 | 1.00 | P<0.0001 | 1.00 | 1.00 | 1.00 | P<0.0001 |

| | | | | | | | | |
|---|---|---|---|---|---|---|---|---|
| 2 | 1.01 | 0.98 | 1.04 | | 1.13 | 1.08 | 1.18 | |
| 3 | 1.04 | 1.01 | 1.07 | | 1.19 | 1.14 | 1.25 | |
| 4 | 1.09 | 1.05 | 1.12 | | 1.31 | 1.25 | 1.38 | |
| 5 (most deprived) | 1.11 | 1.08 | 1.15 | | 1.44 | 1.38 | 1.52 | |
| Smoking history | | | | P<0.0001 | | | | P<0.0001 |
| | 1.05 | 1.03 | 1.07 | | 1.14 | 1.10 | 1.18 | |

*Analyses were controlled for cancer site, patient age, gender, deprivation quintile, year of diagnosis, and history of smoking

**Table A.8. Sensitivity analysis: Associations between emergency presentation route to diagnosis, urgent (two-week-wait) referral route to diagnosis, and patient characteristics (ACG System). ***

| | **Emergency presentation** | | | | **Urgent referral** | | | |
|---|---|---|---|---|---|---|---|---|
| | Odds ratio | Lower 95% confidence interval | Upper 95% confidence interval | P | Odds ratio | Lower 95% confidence interval | Upper 95% confidence interval | P |
| *Morbidity* | | | | | | | | |
| Resource utilisation band 0 | 1.00 | 1.00 | 1.00 | P<0.0001 | 1.00 | 1.00 | 1.00 | P<0.0001 |
| Resource utilisation band 1 | 0.90 | 0.82 | 0.98 | | 0.99 | 0.92 | 1.06 | |
| Resource utilisation band 2 | 0.76 | 0.70 | 0.83 | | 1.00 | 0.94 | 1.07 | |
| Resource utilisation band 3 | 0.75 | 0.69 | 0.81 | | 0.92 | 0.87 | 0.99 | |
| Resource utilisation band 4 | 0.87 | 0.80 | 0.94 | | 0.76 | 0.71 | 0.81 | |
| Resource utilisation band 5 | 1.23 | 1.13 | 1.35 | | 0.56 | 0.53 | 0.61 | |
| Gender (Female) | 1.02 | 1.00 | 1.05 | 0.0384 | 0.97 | 0.95 | 0.99 | 0.0062 |
| *Age* | | | | | | | | |
| 40 to 44 years | 0.84 | 0.77 | 0.91 | P<0.0001 | 0.79 | 0.75 | 0.84 | P<0.0001 |
| 45 to 49 years | 0.90 | 0.85 | 0.96 | | 0.83 | 0.79 | 0.87 | |
| 50 to 54 years | 0.90 | 0.85 | 0.95 | | 0.84 | 0.81 | 0.88 | |
| 55 to 59 years | 0.92 | 0.88 | 0.97 | | 0.91 | 0.87 | 0.94 | |
| 60 to 64 years | 0.95 | 0.91 | 0.99 | | 0.96 | 0.93 | 0.99 | |
| 65 to 69 years | 1.00 | 1.00 | 1.00 | | 1.00 | 1.00 | 1.00 | |
| 70 to 74 years | 1.16 | 1.11 | 1.20 | | 1.00 | 0.97 | 1.03 | |
| 75 to 79 years | 1.35 | 1.30 | 1.40 | | 1.04 | 1.01 | 1.08 | |
| 80 to 84 years | 1.82 | 1.75 | 1.89 | | 0.91 | 0.88 | 0.94 | |
| 85 to 89 years | 2.75 | 2.64 | 2.87 | | 0.74 | 0.71 | 0.77 | |
| 90 years and older | 4.55 | 4.32 | 4.80 | | 0.49 | 0.46 | 0.51 | |
| *IMD* | | | | | | | | |
| 1 (least deprived | 1.00 | 1.00 | 1.00 | P<0.0001 | 1.00 | 1.00 | 1.00 | P<0.0001 |
| 2 | 1.09 | 1.06 | 1.13 | | 1.06 | 1.03 | 1.10 | |
| 3 | 1.18 | 1.14 | 1.22 | | 1.10 | 1.06 | 1.13 | |
| 4 | 1.30 | 1.26 | 1.35 | | 1.06 | 1.02 | 1.10 | |
| 5 (most deprived) | 1.45 | 1.40 | 1.51 | | 1.04 | 1.00 | 1.08 | |
| Smoking history | | | | 0.0047 | | | | P<0.0001 |
| | 1.03 | 1.01 | 1.06 | | 1.08 | 1.06 | 1.11 | |

*Analyses were controlled for cancer site, patient age, gender, deprivation quintile, year of diagnosis, and history of smoking

**Table A.9. Sensitivity analysis: Associations between cancer stage, 30-day mortality, and patient characteristics (morbidity count based on CMS). ***

| | **Advanced-stage cancer diagnosis** | | | | **30-day mortality** | | | |
|---|---|---|---|---|---|---|---|---|
| | Odds ratio | Lower 95% confidence interval | Upper 95% confidence interval | P | Odds ratio | Lower 95% confidence interval | Upper 95% confidence interval | P |
| *Morbidity count* | | | | | | | | |
| 0 | 1.00 | 1.00 | 1.00 | P<0.0001 | 1.00 | 1.00 | 1.00 | P<0.001 |
| 1 | 0.88 | 0.85 | 0.91 | | 0.91 | 0.85 | 0.96 | |
| 2 | 0.82 | 0.79 | 0.84 | | 0.94 | 0.89 | 1.00 | |
| 3 | 0.79 | 0.76 | 0.82 | | 0.99 | 0.93 | 1.05 | |
| 4 or more | 0.69 | 0.67 | 0.72 | | 1.06 | 1.00 | 1.12 | |
| Gender (Female) | 0.88 | 0.85 | 0.90 | P<0.0001 | 0.91 | 0.89 | 0.94 | P<0.0001 |
| *Age* | | | | | | | | |
| 40 to 44 years | 0.64 | 0.59 | 0.68 | P<0.0001 | 0.31 | 0.25 | 0.38 | P<0.0001 |
| 45 to 49 years | 0.76 | 0.72 | 0.80 | | 0.44 | 0.39 | 0.50 | |
| 50 to 54 years | 0.84 | 0.80 | 0.88 | | 0.55 | 0.49 | 0.60 | |
| 55 to 59 years | 0.87 | 0.84 | 0.91 | | 0.66 | 0.61 | 0.71 | |
| 60 to 64 years | 0.94 | 0.91 | 0.97 | | 0.83 | 0.78 | 0.89 | |
| 65 to 69 years | 1.00 | 1.00 | 1.00 | | 1.00 | 1.00 | 1.00 | |
| 70 to 74 years | 1.05 | 1.02 | 1.08 | | 1.26 | 1.20 | 1.34 | |
| 75 to 79 years | 1.12 | 1.08 | 1.16 | | 1.51 | 1.43 | 1.60 | |
| 80 to 84 years | 1.23 | 1.18 | 1.28 | | 2.20 | 2.09 | 2.33 | |
| 85 to 89 years | 1.41 | 1.35 | 1.48 | | 3.32 | 3.13 | 3.53 | |
| 90 years and older | 1.56 | 1.45 | 1.67 | | 6.00 | 5.61 | 6.42 | |
| *IMD* | | | | | | | | |
| 1 (least deprived | 1.00 | 1.00 | 1.00 | P<0.0001 | 1.00 | 1.00 | 1.00 | P<0.0001 |
| 2 | 1.02 | 0.99 | 1.05 | | 1.13 | 1.08 | 1.18 | |
| 3 | 1.05 | 1.01 | 1.08 | | 1.19 | 1.14 | 1.25 | |
| 4 | 1.10 | 1.06 | 1.13 | | 1.31 | 1.25 | 1.38 | |
| 5 (most deprived) | 1.13 | 1.09 | 1.16 | | 1.45 | 1.38 | 1.52 | |
| Smoking history | 1.06 | 1.03 | 1.08 | P<0.0001 | 1.14 | 1.10 | 1.18 | P<0.0001 |

*Analyses were controlled for cancer site, patient age, gender, deprivation quintile, year of diagnosis, and history of smoking

**Table A.10. Sensitivity analysis: Associations between emergency presentation route to diagnosis, urgent (two-week-wait) referral route to diagnosis, and patient characteristics (morbidity count based on CMS). ***

| | **Emergency presentation** | | | | **Urgent suspected cancer referral** | | | |
|---|---|---|---|---|---|---|---|---|
| | Odds ratio | Lower 95% confidence interval | Upper 95% confidence interval | P | Odds ratio | Lower 95% confidence interval | Upper 95% confidence interval | P |
| *Morbidity count* | | | | | | | | |
| 0 | 1.00 | 1.00 | 1.00 | P<0.0001 | 1.00 | 1.00 | 1.00 | P<0.0001 |
| 1 | 0.93 | 0.90 | 0.97 | | 0.98 | 0.95 | 1.01 | |
| 2 | 0.94 | 0.90 | 0.97 | | 0.93 | 0.91 | 0.96 | |
| 3 | 0.95 | 0.91 | 0.99 | | 0.89 | 0.86 | 0.92 | |
| 4 or more | 1.08 | 1.04 | 1.12 | | 0.77 | 0.75 | 0.79 | |
| Gender (Female) | 1.01 | 0.99 | 1.04 | 0.2068 | 0.97 | 0.95 | 1.00 | 0.0175 |
| *Age* | | | | | | | | |
| 40 to 44 years | 0.86 | 0.80 | 0.94 | P<0.0001 | 0.77 | 0.73 | 0.82 | P<0.0001 |
| 45 to 49 years | 0.93 | 0.87 | 0.98 | | 0.82 | 0.78 | 0.86 | |
| 50 to 54 years | 0.92 | 0.87 | 0.97 | | 0.83 | 0.80 | 0.86 | |
| 55 to 59 years | 0.94 | 0.89 | 0.98 | | 0.89 | 0.86 | 0.93 | |
| 60 to 64 years | 0.96 | 0.92 | 1.00 | | 0.95 | 0.92 | 0.99 | |
| 65 to 69 years | 1.00 | 1.00 | 1.00 | | 1.00 | 1.00 | 1.00 | |
| 70 to 74 years | 1.15 | 1.11 | 1.20 | | 1.01 | 0.98 | 1.04 | |
| 75 to 79 years | 1.34 | 1.29 | 1.39 | | 1.05 | 1.02 | 1.09 | |
| 80 to 84 years | 1.81 | 1.74 | 1.88 | | 0.92 | 0.89 | 0.95 | |

| | | | | | | | | |
|---|---|---|---|---|---|---|---|---|
| 85 to 89 years | 2.75 | 2.63 | 2.87 | | 0.74 | 0.71 | 0.77 | |
| 90 years and older | 4.56 | 4.33 | 4.81 | | 0.49 | 0.46 | 0.51 | |
| *IMD* | | | | | | | | |
| 1 (least deprived | 1.00 | 1.00 | 1.00 | P<0.0001 | 1.00 | 1.00 | 1.00 | P<0.0001 |
| 2 | 1.09 | 1.06 | 1.13 | | 1.07 | 1.04 | 1.10 | |
| 3 | 1.18 | 1.14 | 1.22 | | 1.10 | 1.06 | 1.13 | |
| 4 | 1.30 | 1.26 | 1.35 | | 1.06 | 1.03 | 1.10 | |
| 5 (most deprived) | 1.45 | 1.40 | 1.51 | | 1.04 | 1.00 | 1.08 | |
| Smoking history | | | | 0.0040 | | | | P<0.0001 |
| | 1.04 | 1.01 | 1.06 | | 1.08 | 1.06 | 1.11 | |

*Analyses were controlled for cancer site, patient age, gender, deprivation quintile, year of diagnosis, and history of smoking

### Figures

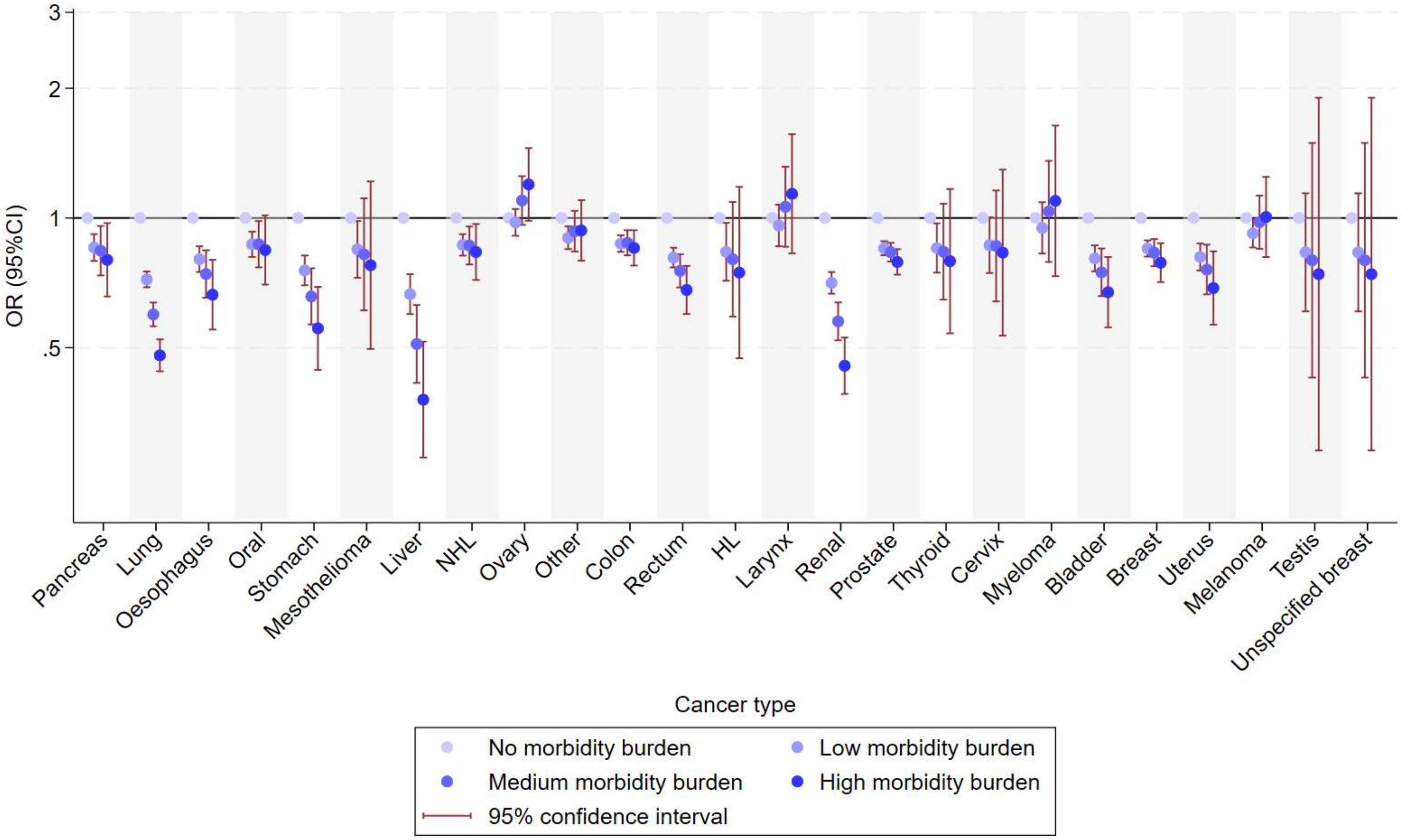

**Figure. A.1** Associations between cancer stage and morbidity burden by cancer type

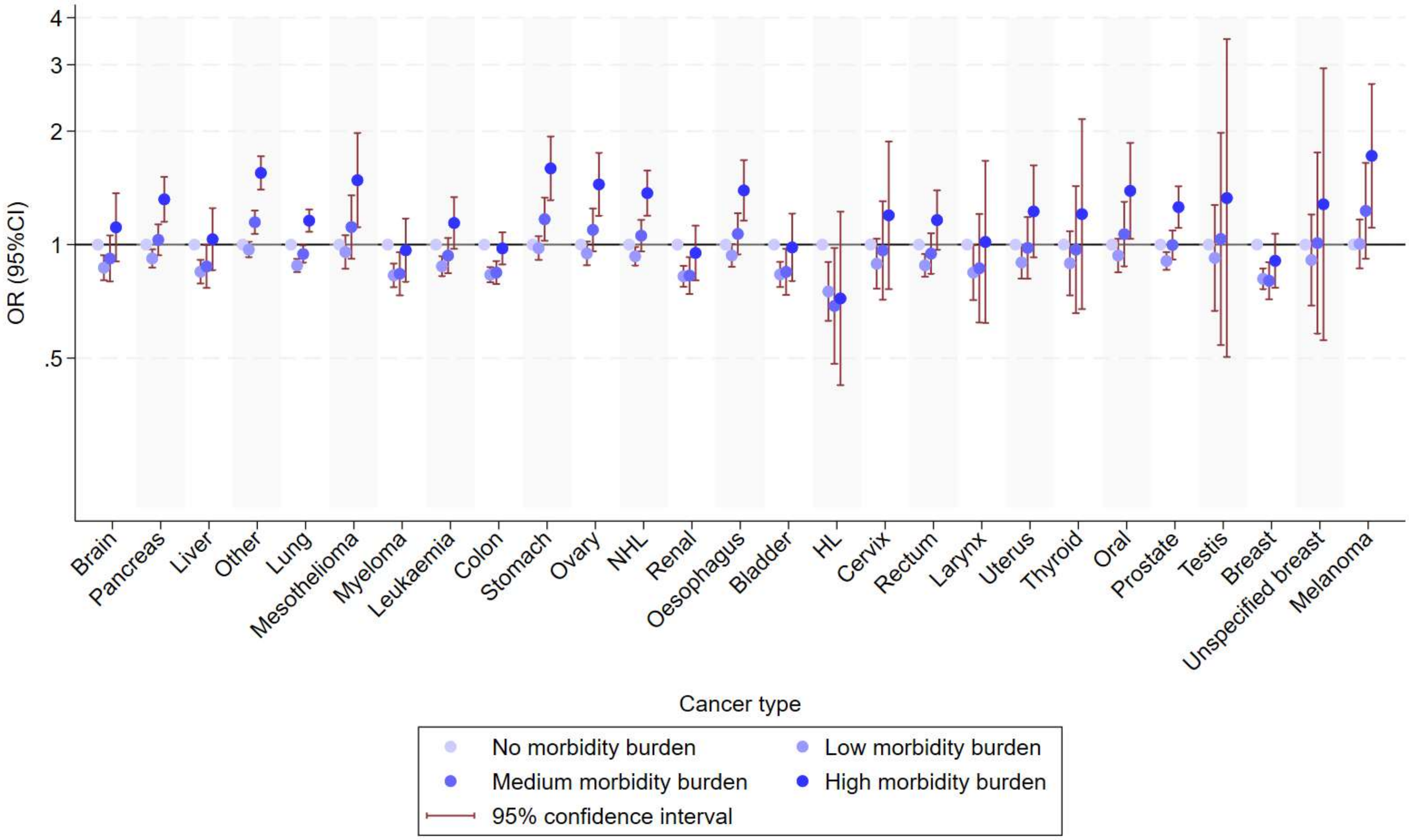

**Figure. A.2** Associations between emergency presentation route to diagnosis and morbidity burden by cancer type

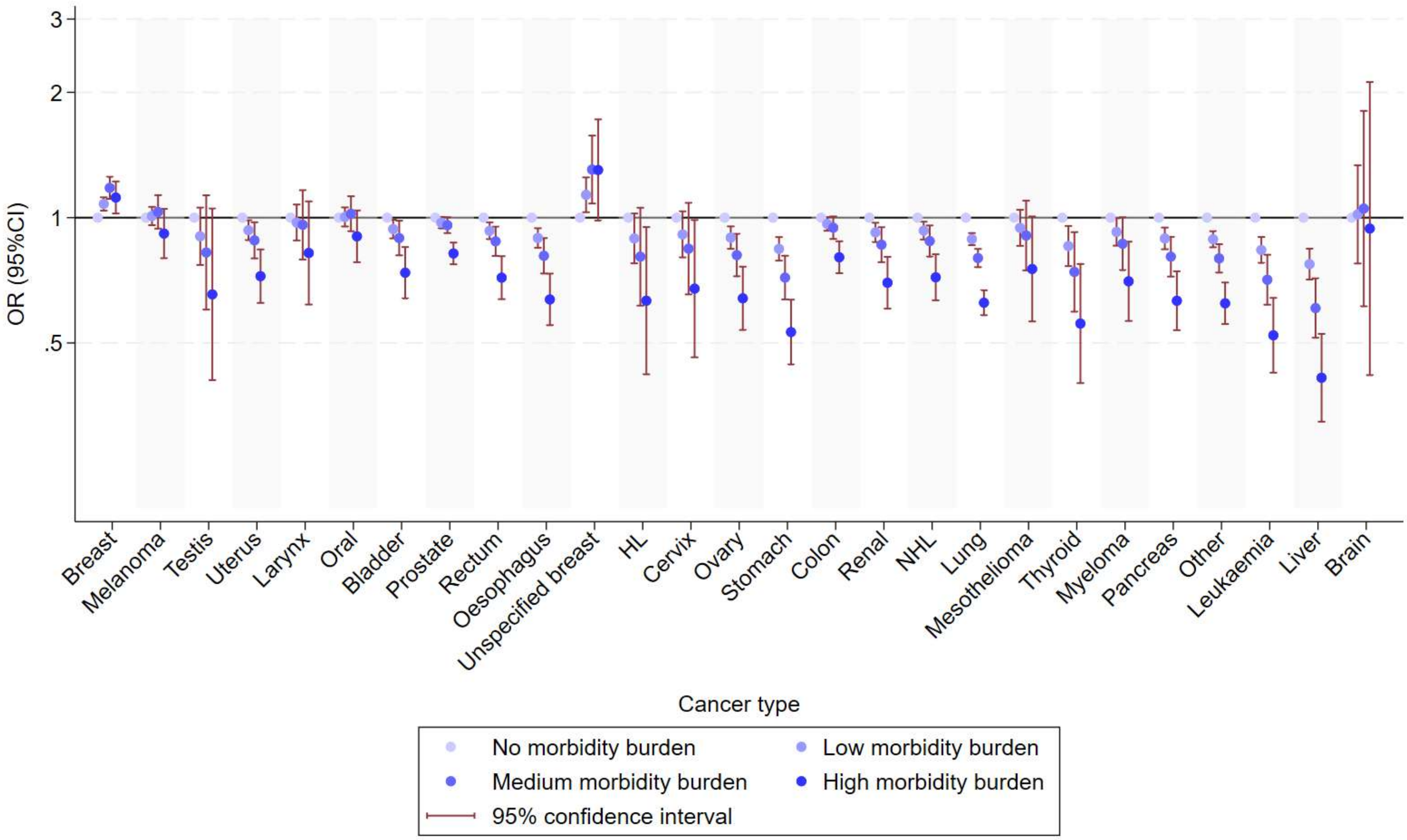

**Figure. A.3** Associations between an urgent suspected cancer referral route to diagnosis and morbidity burden by cancer type

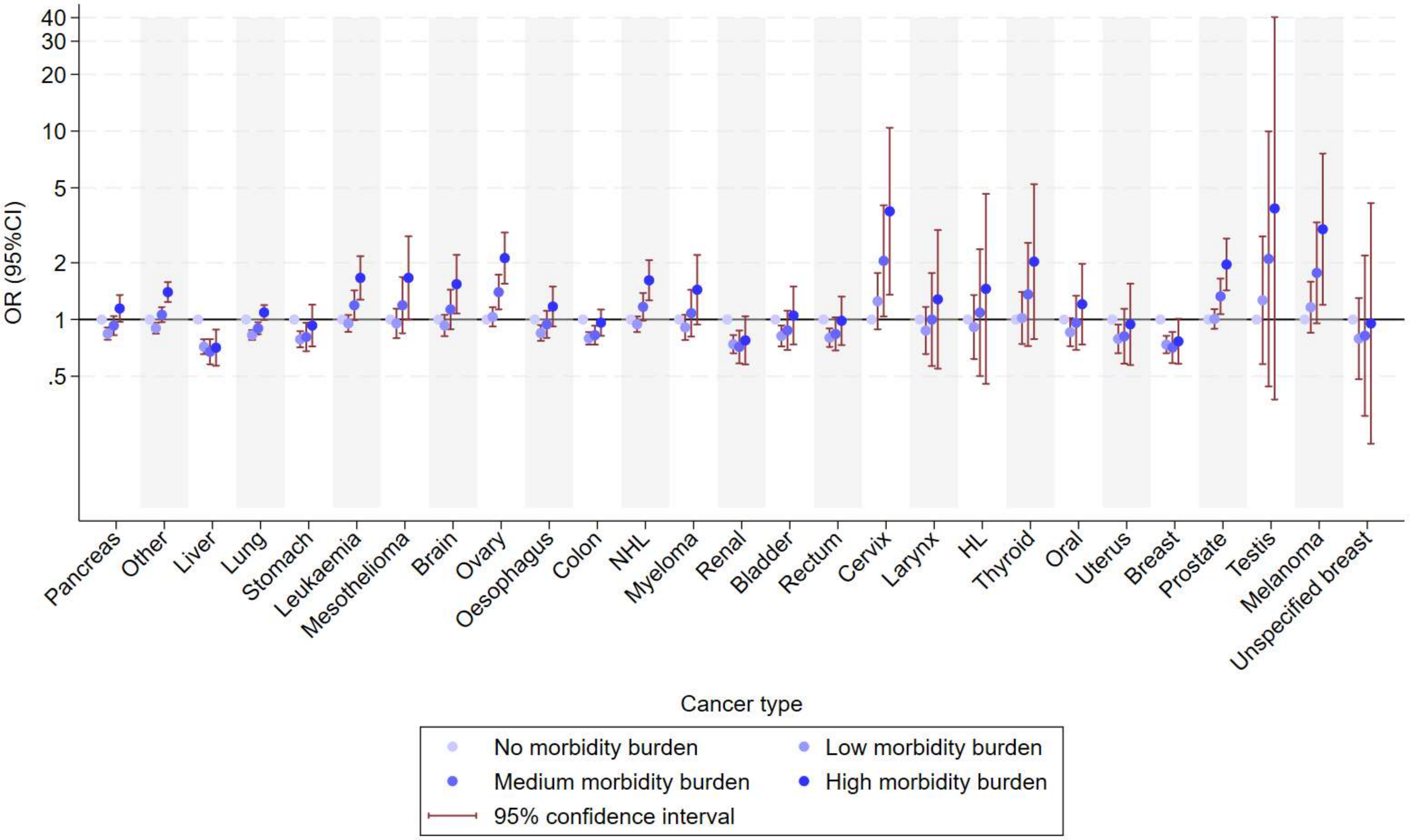

**Figure. A.4** Associations between 30-day mortality and morbidity burden by cancer type
